# Game Over for the Baseline: Anomalous Burden and Structural Seasonal Shifts in Post-Pandemic U.S. Influenza Hospitalization, 2009–2025

**DOI:** 10.64898/2026.03.15.26348430

**Authors:** Hayden D. Hedman

## Abstract

**Background/Objectives:** The trajectory of influenza hospitalization burden from pre-pandemic baseline through post-pandemic recovery remains poorly characterized at the national level. This study characterized phase-stratified burden and seasonal structure, quantified racial and ethnic disparities, and assessed whether post-pandemic seasons represent anomalous departures from pre-pandemic expectations.

**Methods:** Sixteen seasons of FluSurv-NET surveillance data (2009–2010 through 2024–2025; 509 observation weeks) were analyzed across pre-pandemic, disruption, and recovery phases using OLS regression with effect-size estimation, bootstrapped age-adjusted rate ratios, seasonal-trend decomposition (STL), Prophet time-series forecasting, and Isolation Forest anomaly detection.

**Results:** Mean peak weekly hospitalization rate nearly doubled from pre-pandemic to recovery (5.1 to 11.1 per 100,000), cumulative seasonal burden increased from 46.3 to 87.0 per 100,000, and median peak timing advanced from MMWR week 9 to week 50. STL decomposition revealed a marked shift from weak pre-pandemic seasonality (Fs = 0.14) to substantially stronger annual regularity (Fs = 0.98) across three recovery seasons, with threefold amplitude increase. Non-Hispanic Black persons had rate ratios of 1.72, 2.16, and 1.99 relative to White persons across phases; American Indian and Alaska Native persons showed the highest disruption-phase ratio (2.24, 95% CI 1.90–3.53), based on two contributing seasons. A flat-growth Prophet model detected first exceedance in February 2020, outperforming a linear-growth specification on held-out validation. Isolation Forest identified 2017–2018, 2023–2024, and 2024–2025 as robust anomalies across all contamination thresholds.

**Conclusions:** Post-pandemic influenza recovery is characterized by intensified and restructured seasonality, persistent racial and ethnic disparities, and anomalous burden exceeding pre-pandemic projections, identified independently by time-series forecasting and unsupervised anomaly detection.

## 1. Introduction

Seasonal influenza remains a substantial cause of morbidity, hospitalization, and mortality in the United States, with burden concentrated among adults aged 65 years and older, young children, and individuals with underlying medical conditions [1,2]. The Influenza Hospitalization Surveillance Network (FluSurv-NET) [3]: a population-based platform covering approximately 10% of the US population across 14 states, has provided the primary infrastructure for tracking laboratory-confirmed influenza hospitalization rates since the 2009-2010 pandemic season [3]. Despite this surveillance infrastructure, the full epidemiological trajectory from the pre-pandemic baseline through pandemic disruption and into the post-pandemic recovery period has not been comprehensively characterized at the national level using the complete FluSurv-NET time series.

The COVID-19 pandemic profoundly disrupted influenza transmission dynamics. Non-pharmaceutical interventions implemented beginning in 2020 suppressed influenza circulation to near-zero levels during the 2020-2021 season, with an extended period of minimal transmission giving rise to what has been termed immunological debt: a population-level reduction in naturally acquired immunity due to the absence of antigenic exposure over multiple seasons, resulting in an expanded susceptible pool when transmission resumed [4–6]. Evidence for this phenomenon has been documented across multiple respiratory pathogens in the post-pandemic period [5–7], though data specifically characterizing the magnitude, trajectory, and structural character of influenza’s post-pandemic recovery in the United States remain limited [8,9]. The 2022-2023 through 2024-2025 seasons represent the first complete recovery-phase influenza seasons available for systematic longitudinal analysis.

Racial and ethnic disparities in influenza hospitalization burden have been documented persistently across seasons, with non-Hispanic Black and American Indian and Alaska Native populations consistently experiencing higher hospitalization rates than non-Hispanic White populations [10,11]. Whether the disruption and recovery periods widened or attenuated pre-existing disparities remains an open empirical question

Existing analyses of post-pandemic influenza dynamics have largely focused on single seasons, specific subpopulations, or burden estimation methodologies rather than longitudinal characterization of the full pre-pandemic through recovery trajectory [7,9]. Longitudinal characterization of influenza hospitalization burden and seasonal structure across the full pandemic transition remains incomplete, and multi-method analytic approaches applied to population-based surveillance data spanning this period are limited.

To address this gap, longitudinal analysis of FluSurv-NET surveillance data was applied to examine: (1) phase-stratified influenza hospitalization burden and seasonal structure across the pre-pandemic, disruption, and recovery periods; (2) racial and ethnic disparities in hospitalization rates and their evolution across phases; and (3) whether recovery-phase seasons represent anomalous departures from pre-pandemic expectations. Post-pandemic influenza recovery was characterized by compressed but intensified seasonality, persistent and widened racial and ethnic disparities, and anomalous peak burden in the two most recent complete seasons exceeding pre-pandemic projections across multiple independent analytic methods.

## 2. Materials and Methods

### 2.1. Study design and data source

Population-based longitudinal surveillance data were obtained from the Influenza Hospitalization Surveillance Network (FluSurv-NET), operated by the Centers for Disease Control and Prevention (CDC) as a component of the Respiratory Virus Hospitalization Surveillance Network (RESP-NET) [3]. FluSurv-NET conducts active, laboratory-confirmed influenza hospitalization surveillance across more than 90 counties in 14 states, representing approximately 34 million persons and an estimated 10% of the U.S. population across nine Department of Health and Human Services regions [3].

A case was defined as a resident of a FluSurv-NET catchment area who received a positive influenza laboratory test ordered by a healthcare professional within 14 days before or during hospitalization. Acceptable laboratory methods included viral culture, direct or indirect fluorescent antibody staining, rapid antigen testing, and molecular assay [3,12]. No additional inclusion or exclusion criteria were applied beyond CDC’s published case definition and suppression rules. Data were extracted from FluView Interactive on 7 March 2026 [3]. All analyses were restricted to the Entire Network catchment stratum to ensure geographic comparability across seasons; sub-network and state-level strata were excluded to avoid differential catchment effects introduced by changes in network composition over time.

### 2.2. Study period and phase classification

The analytic dataset spanned the 2009–2010 through 2024–2025 influenza seasons, comprising 16 seasons of population-based surveillance. The in-progress 2025–2026 season was retained for descriptive summaries but excluded from all time-series models and regression analyses requiring complete seasonal data.

Seasons were classified into three a priori epidemiological phases. The pre-pandemic baseline phase comprised the 2009–2010 through 2018–2019 seasons (ten seasons), representing the normative range of influenza hospitalization burden prior to SARS-CoV-2 emergence. The pandemic disruption phase comprised the 2019–2020 through 2021–2022 seasons (three seasons), characterized by suppressed influenza circulation attributable to non-pharmaceutical interventions, behavioral change, and competitive viral interference during the COVID-19 pandemic [13–15]. The post-pandemic recovery phase comprised the 2022–2023 season onward (four complete seasons), reflecting re-establishment of seasonal influenza dynamics following relaxation of pandemic-era measures.

The 2009–2010 influenza A(H1N1) pandemic season was retained in all primary analyses but flagged as a binary covariate in sensitivity models to account for its atypical epidemiological profile [12]. Weekly hospitalization rates for the 2020–2021 season were suppressed by CDC due to low counts and data quality concerns arising from COVID-19 cocirculation; this season contributed 31 observation weeks with non-null cumulative rates but null weekly rates. These observations were retained as confirmed structural absences and were not imputed; rate-based analyses for 2020–2021 relied exclusively on cumulative rates.

### 2.3. Surveillance sampling and denominator considerations

Two methodological discontinuities require consideration for valid inference across the full study period. Beginning with the 2017–2018 season, FluSurv-NET implemented probability-based sampling for clinical data collection: patients aged 50 and older were age-stratified and sampled below 100%, while all in-hospital deaths were captured in full regardless of age *[3]*. Clinical data elements subject to this design should be interpreted as weighted estimates rather than complete case counts.

A second discontinuity affects population denominators used to compute hospitalization rates: seasons prior to 2020–2021 used National Center for Health Statistics bridged-race population estimates, while seasons from 2020–2021 onward used unbridged U.S. Census Bureau estimates following discontinuation of bridged-race products *[3]*. This transition introduces a comparability limitation for absolute rate comparisons across the disruption boundary; within-phase comparisons are internally consistent. Cross-phase absolute rate comparisons were examined with a denominator-version indicator variable included in sensitivity models.

### 2.4. Data Management and Variable Construction

Raw surveillance data were preprocessed to address suppression artifacts, temporal indexing, and season-boundary ambiguities inherent to multi-season public health surveillance extracts. CDC suppression markers were replaced with missing values; confidence interval fields were absent across all strata and excluded from analysis. Season-boundary weeks appearing in two consecutive seasons were resolved by retaining the higher weekly rate to preserve peak burden estimates [3]. A continuous weekly time series was constructed using MMWR epiweeks. The 2020–2021 season was confirmed to reflect genuine near-zero influenza activity rather than data loss, based on non-null cumulative rate records [3].

All time-series and phase-level inference used the overall network stratum. Stratified subsets by age group, sex, race/ethnicity, and virus type were used for descriptive and disparity analyses. All analyses were conducted in Python 3.11 using pandas, NumPy, statsmodels, scikit-learn, and Prophet. A fixed random seed was applied to all stochastic procedures to ensure reproducibility (SM1).

### 2.5. Descriptive Analysis

Descriptive summaries were computed across five stratification dimensions: overall network, five standard CDC age groups (0–4, 5–17, 18–49, 50–64, and ≥65 years), sex (male and female), race/ethnicity (White, Black, Hispanic/Latino, Asian/Pacific Islander, and American Indian/Alaska Native), and influenza virus type (Influenza A, Influenza B, A(H1N1)pdm09, and A(H3N2)) [3].

The variance-to-mean ratio of weekly hospitalization rates was computed within each stratum prior to summarization to evaluate distributional properties and inform the choice of central tendency measure [16]. Median with interquartile range was used as the primary phase-level summary, with mean and standard deviation as secondary estimates. Season-level metrics were computed first for each stratum and phase, including peak weekly rate, peak MMWR week, end-of-season cumulative rate, and active season duration, defined as the number of weeks with a weekly rate ≥0.1 per 100,000 population. These season-level values were then averaged within phase to prevent seasons with longer observation windows from exerting disproportionate influence on phase estimates. The 2020–2021 season was excluded from weekly rate summaries but retained in cumulative rate summaries.

### 2.6. Race/ethnicity disparity analysis

Age-adjusted cumulative hospitalization rate ratios were estimated by race/ethnicity and epidemiological phase using the White population as the reference group. The outcome for each season was the end-of-season maximum age-adjusted cumulative hospitalization rate. Phase-level rate ratios were calculated as the ratio of phase means across seasons, with 95% confidence intervals derived by nonparametric bootstrap resampling of season-level pairs (1,000 iterations, percentile method). The 2020–2021 season was excluded because all race-stratified weekly rates were structurally suppressed. Cross-phase comparisons of absolute rate levels were interpreted with caution given the denominator discontinuity described in Section 2.3; within-phase rate ratios are the primary inferential targets.

### 2.7. Phase-level regression analysis

The overall-stratum weekly rate series was aggregated to the season level, yielding one mean weekly rate per season, and regressed on phase dummy variables using ordinary least squares with the pre-pandemic phase as the reference category. The analytic sample comprised 15 seasons after excluding 2020–2021 and the partial 2025–2026 season. A sensitivity model additionally adjusted for denominator version and the 2009–2010 pandemic indicator. Effect sizes were quantified as Cohen’s d computed pairwise between phases using pooled standard deviation [17]. The gap between unadjusted and adjusted R^2^ in the primary model reflects limited degrees of freedom at n = 15 seasons; effect size estimation is foregrounded accordingly.

### 2.8. Linear mixed-effects model

A linear mixed-effects model was fit to weekly hospitalization rates with within-season week number as a fixed effect and a season-level random intercept, estimated by restricted maximum likelihood. Phase was not included as a fixed effect, as phase classification is invariant within season and would be fully absorbed by the season-level random intercept. The within-season week number coefficient is interpreted as the average within-season rate trajectory across all modeled seasons, independent of between-season burden differences captured by the random intercept.

### 2.9. Seasonal decomposition

Seasonal-trend decomposition using LOESS (STL) was applied separately to the pre-pandemic series (2009–2010 through 2018–2019; 300 weeks) and the post-pandemic recovery series (2022–2023 through 2024–2025; 112 weeks) to isolate structural changes in seasonality independent of trend [18]. The pandemic disruption phase was excluded from both fits. Decomposition used a period of 52 weeks with robust LOESS fitting. Seasonal amplitude was quantified as the interquartile range of the seasonal component. Strength of seasonality (Fs) and strength of trend (Ft) were computed [19]. Post-pandemic series estimates should be interpreted with the constraint of three contributing seasons; the consistency of directional shifts across all four decomposition metrics supports the observed structural change.

### 2.10. Time-series forecasting and anomaly detection

A Prophet time-series model was fit to the overall-stratum weekly rate series using all non-null pre-pandemic weeks as training data (300 weeks; 27 September 2009 to 21 April 2019) to generate out-of-sample forecasts representing expected rates under pre-pandemic seasonal dynamics [20,21]. The primary model specified flat growth with additive yearly seasonality, a changepoint prior scale of 0.05, 15 potential changepoints, and 95% prediction intervals derived from 1,000 uncertainty samples. A flat growth specification was selected over linear growth to avoid extrapolating the pre-pandemic upward trend into the forecast period, which would inflate projected baselines and delay anomaly detection. A sensitivity model substituting linear growth was fit to the same training data. Both models were evaluated on a held-out validation season (2018–2019) withheld prior to out-of-sample projection. Detection stability was assessed via leave-one-out refitting, excluding each pre-pandemic season in turn and recording the first exceedance date (see SM4). The first post-training week in which the observed rate exceeded the upper bound of the 95% prediction interval was identified as the anomaly onset date for each model.

### 2.11. Isolation Forest anomaly detection

The Isolation Forest algorithm was applied to a season-level feature matrix constructed from the overall-stratum weekly rate series to identify epidemiologically anomalous seasons without imposing distributional assumptions [22]. Six features were derived for each season: peak weekly rate, mean weekly rate, cumulative rate, number of observed weeks, peak MMWR week, and coefficient of variation of weekly rates. The 2020–2021 season was excluded from the feature matrix due to CDC structural data suppression. Features were standardized using parameters derived from pre-pandemic seasons only, ensuring that the normative distribution reflected pre-pandemic burden. The primary model used 500 trees and a contamination parameter of 0.10. Robustness was assessed via a sensitivity sweep across contamination values of 0.05, 0.10, and 0.15; seasons flagged as anomalous across all three values were designated robust anomalies. Cross-method convergence between Prophet forecast gaps and Isolation Forest classifications was assessed for all seasons and is presented in SM4.

## 3. Results

### 3.1. Phase-stratified descriptive findings

The analytic dataset comprised 509 observation weeks across 16 influenza seasons including one partial season (2024–2025). Overall weekly hospitalization rates were broadly comparable between the pre-pandemic and pandemic disruption phases at the median level (0.8 and 0.6 per 100,000, respectively), while the post-pandemic recovery phase showed both higher median weekly rates (1.0 per 100,000) and substantially elevated peak and cumulative burden. Mean peak weekly rate nearly doubled from 5.1 per 100,000 (SD 2.8) in the pre-pandemic phase to 11.1 per 100,000 (SD 2.7) in recovery, and mean end-of-season cumulative rate increased from 46.3 to 87.0 per 100,000. Season timing shifted markedly, with median peak MMWR week advancing from week 9 in the pre-pandemic phase to week 50 in recovery, reflecting a transition from late-winter to early-winter seasonal peaks. Active season duration remained stable across all three phases (median 30 weeks). Full phase-and season-level descriptive statistics are presented in Table 1 (Figure 1).

**Table 1.**
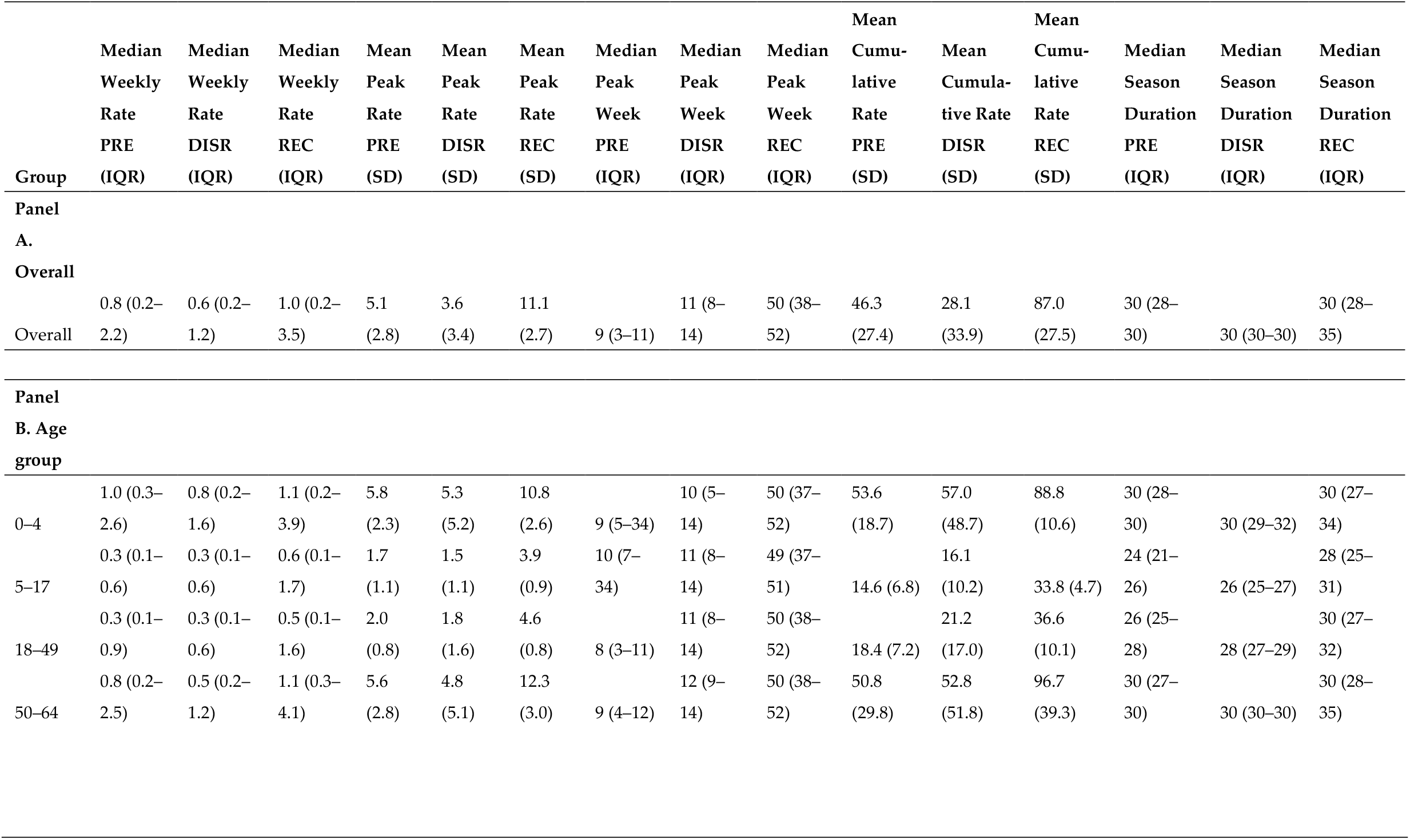
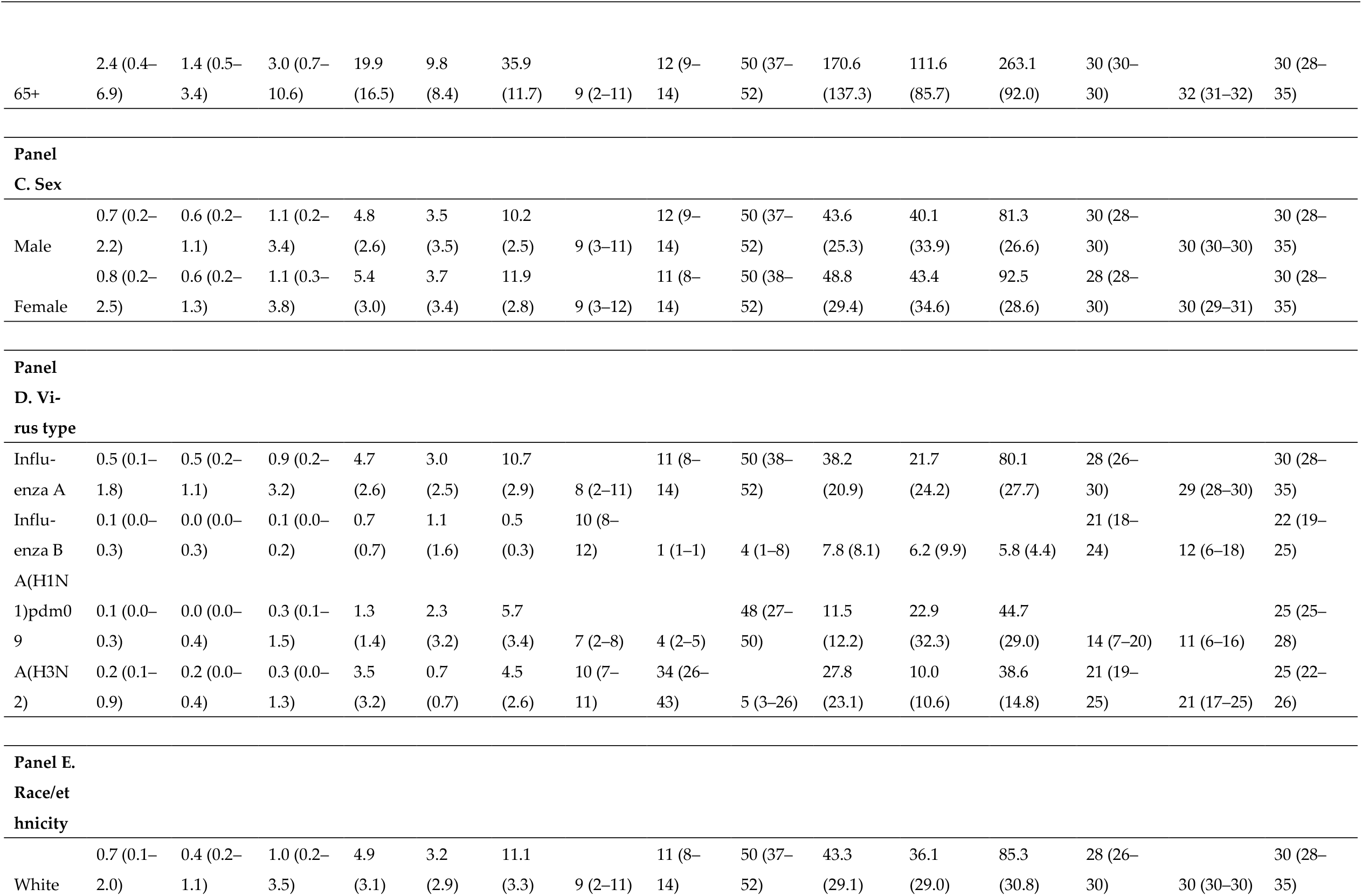
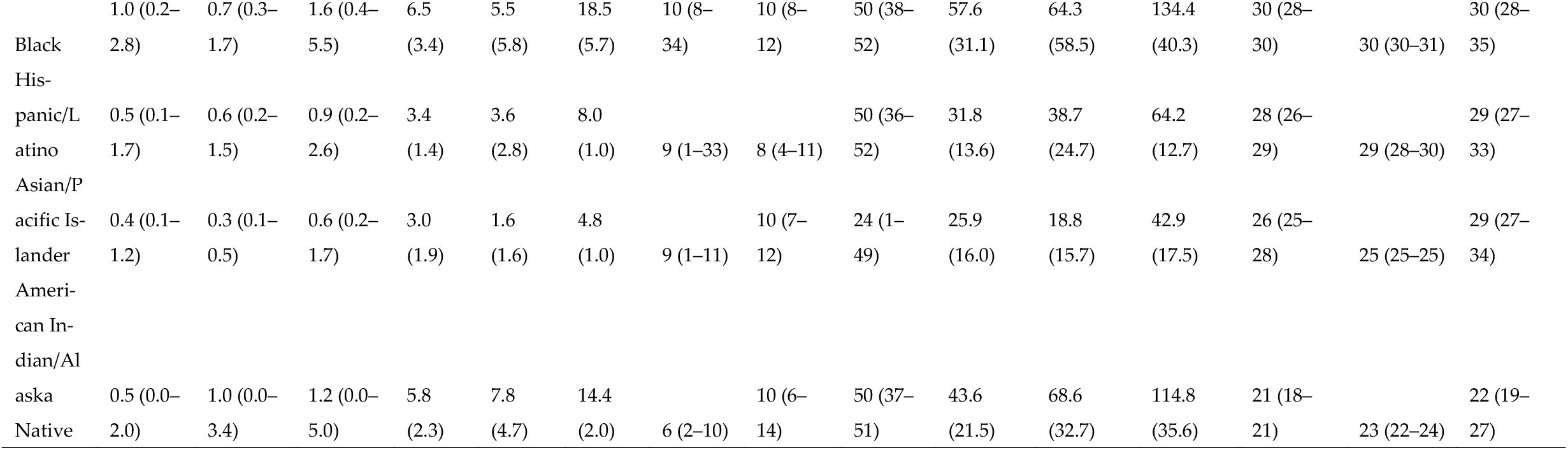
Phase-stratified influenza hospitalization rates by demographic and virologic subgroup, FluSurv-NET, United States, 2009–2025. All rates are per 100,000 population. Phase abbreviations: PRE = pre-pandemic baseline (2009–10 to 2018–19); DISR = pandemic disruption (2019–20 to 2021–22); REC = post-pandemic recovery (2022–23 to 2024–25). Weekly rate summaries exclude the 2020–21 season, during which CDC structurally suppressed weekly hospitalization data; cumulative rates for 2020–21 are retained where available. Peak week is reported as MMWR epidemiological week number. Season duration reflects the number of weeks with a weekly hospitalization rate ≥0.1 per 100,000. IQR = interquartile range; SD = standard deviation; AIAN = American Indian/Alaska Native; API = Asian/Pacific Islander.

**Figure 1.**
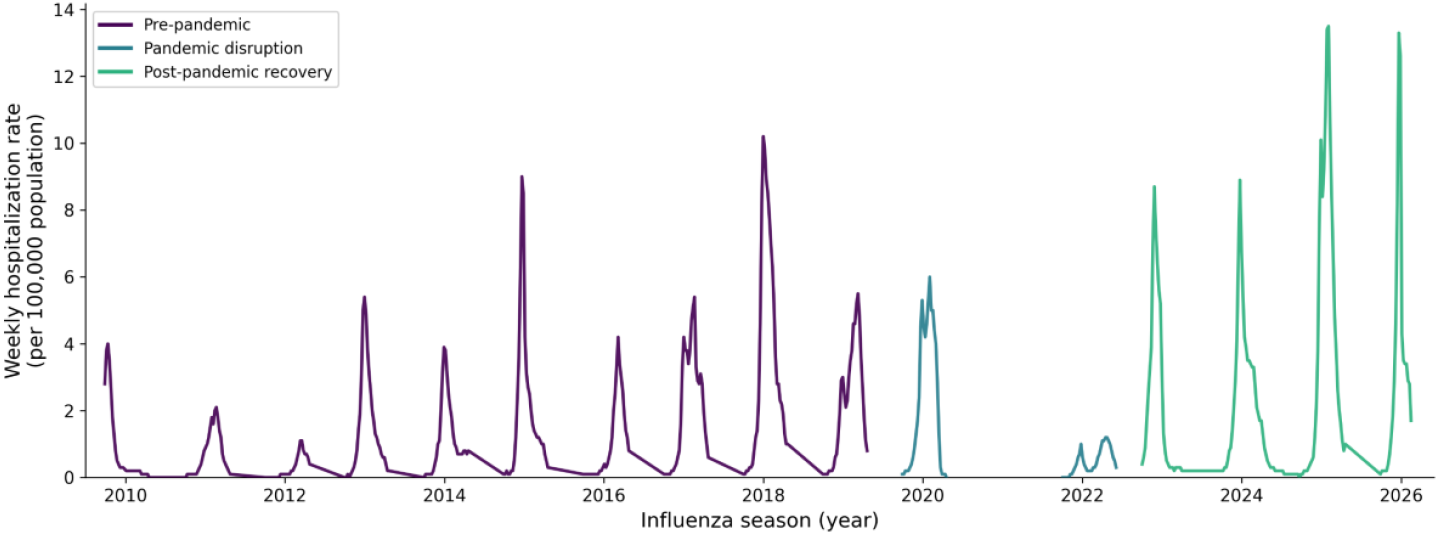
Weekly influenza hospitalization rates per 100,000 population, FluSurv-NET surveillance network, United States, 2009–2025.

By age group, adults aged 65 and older carried the highest burden across all three phases, with mean peak weekly rates of 19.9, 9.8, and 35.9 per 100,000 in the pre-pandemic, disruption, and recovery phases respectively — substantially exceeding all younger groups in every phase (Figure 2). Adults aged 50–64 showed the largest absolute increase in mean cumulative rate from pre-pandemic to recovery (50.8 to 96.7 per 100,000), followed by children aged 0–4 (53.6 to 88.8 per 100,000). The recovery-phase shift in peak timing to early winter was consistent across all age groups. Sex-stratified rates were similar between males and females within each phase, with females showing marginally higher mean cumulative rates in recovery (92.5 vs. 81.3 per 100,000). By virus type, Influenza A dominated all three phases and drove the majority of recovery-phase burden (mean cumulative rate 80.1 per 100,000), while Influenza B remained markedly suppressed through the disruption phase and into recovery (mean cumulative rate 5.8 per 100,000). Both A(H1N1)pdm09 and A(H3N2) subtypes showed recovery-phase resurgence with substantially shifted peak timing relative to pre-pandemic baselines.

**Figure 2.**
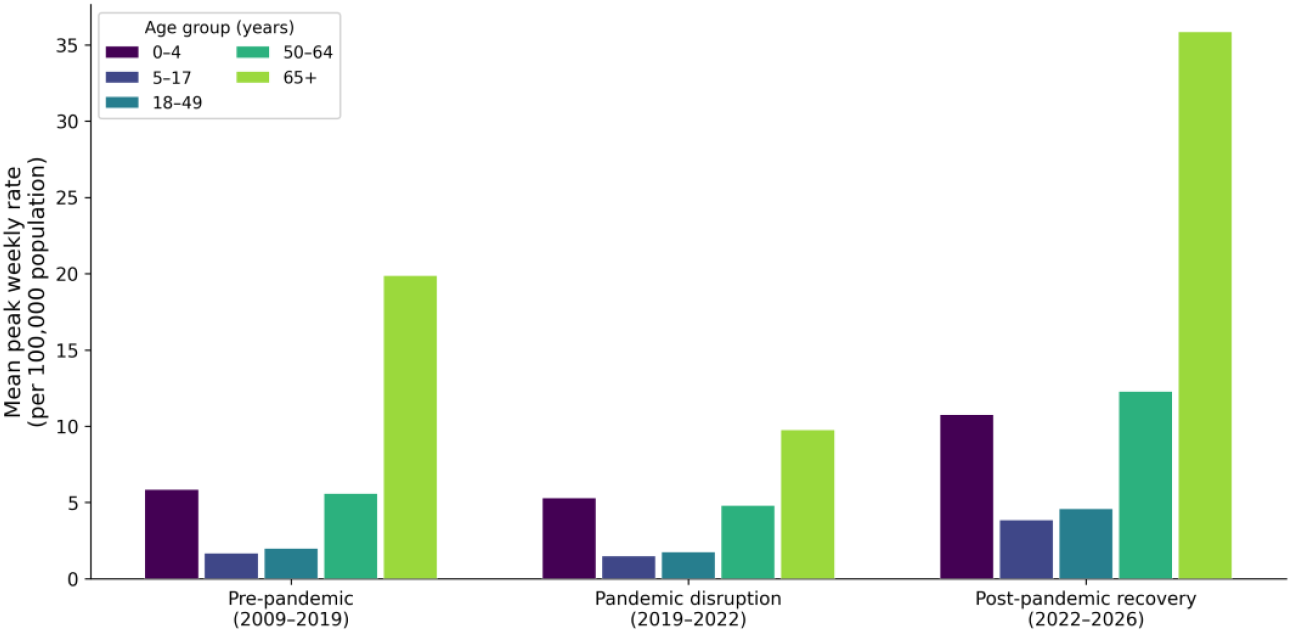
Mean peak weekly influenza hospitalization rates by age group and epidemiological phase, FluSurv-NET surveillance network, United States, 2009–2025.

### 3.2. Race/ethnicity disparities in hospitalization rates

Racial and ethnic disparities in age-adjusted influenza hospitalization rates were present across all three epidemiological phases and persisted into the post-pandemic recovery period (Table 2; Figure 3). Black persons experienced the most consistently elevated burden relative to the White reference group, with rate ratios of 1.72 (95% CI 1.65– 1.83), 2.16 (95% CI 1.75–2.27), and 1.99 (95% CI 1.88–2.17) in the pre-pandemic, disruption, and recovery phases, respectively. The disruption-phase elevation represents a widening of a pre-existing disparity that attenuated only partially in recovery. American Indian/Alaska Native persons had the highest disruption-phase rate ratio of any group examined (RR 2.24, 95% CI 1.90–3.53), though this estimate is based on two seasons with mixed denominator versions and carries substantial uncertainty as reflected in the wide confidence interval; the recovery-phase ratio attenuated to 1.63 (95% CI 1.39–2.00). Hispanic/Latino persons showed a pre-pandemic elevation with confidence intervals spanning the null (RR 1.11, 95% CI 0.99–1.32), a clearly elevated disruption-phase ratio (RR 1.52, 95% CI 1.39–2.00), and a return toward pre-pandemic levels in recovery (RR 1.11, 95% CI 1.00–1.29). Asian/Pacific Islander persons consistently showed rates below the White reference across all phases (RR 0.75, 0.66, and 0.65 in the pre-pandemic, disruption, and recovery phases, respectively); the narrow disruption-phase confidence interval (95% CI 0.61–0.67) reflects limited between-season variability in this group across the two available seasons rather than high estimation precision. Cross-phase comparisons of absolute adjusted rates should be interpreted with caution given the population denominator discontinuity at the 2020–21 season boundary; within-phase rate ratios are internally consistent.

**Table 2.**
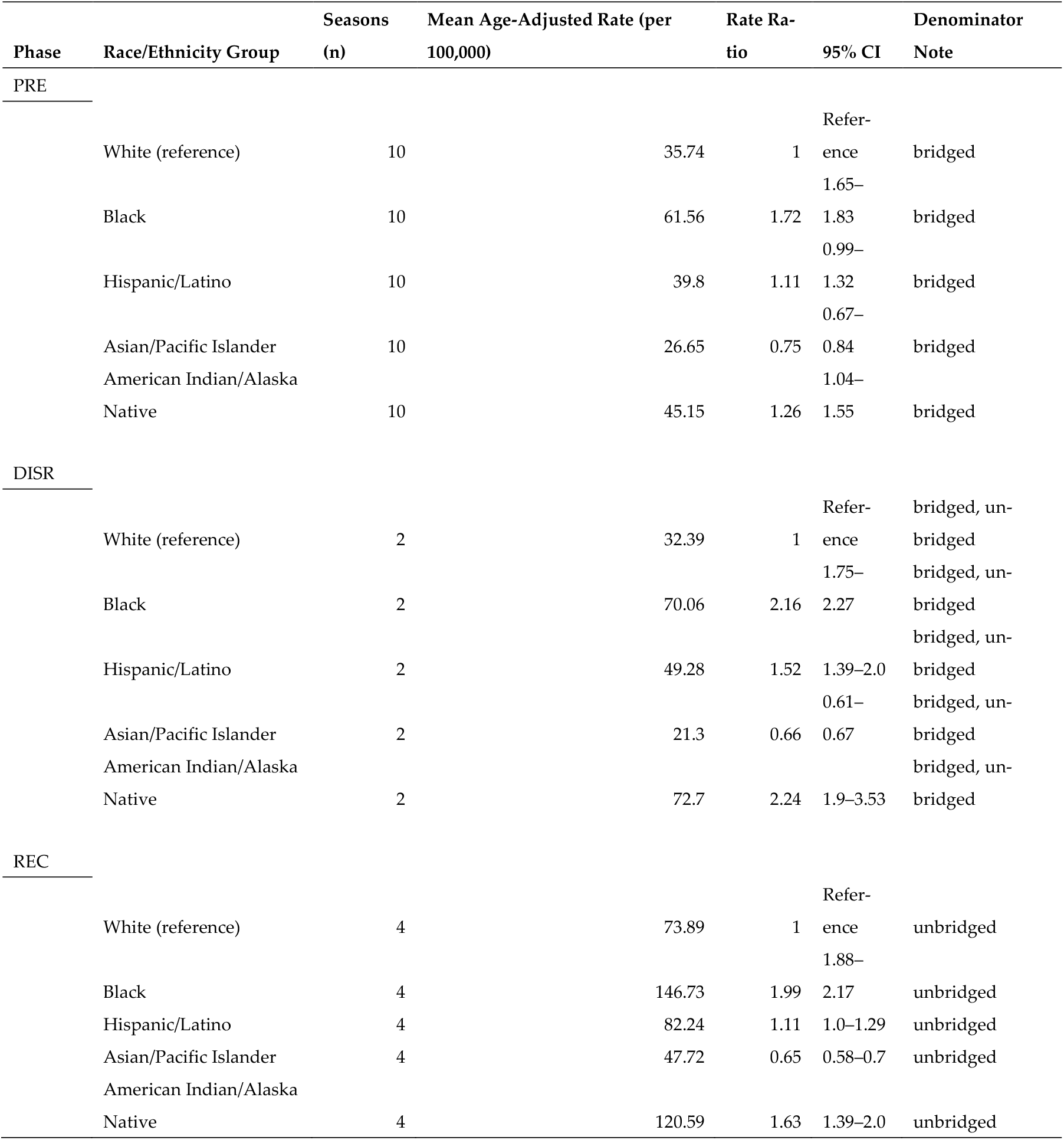
Age-adjusted influenza hospitalization rate ratios by race/ethnicity and epidemiological phase, FluSurv-NET, United States, 2009–2025. Rate ratios compare age-adjusted end-of-season cumulative hospitalization rates for each racial/ethnic group relative to the Non-Hispanic White reference group. Phase abbreviations: PRE = pre-pandemic baseline (2009–10 to 2018–19); DISR = pandemic disruption (2019–20 to 2021–22); REC = post-pandemic recovery (2022–23 to 2024–25). The 2020–21 season was excluded from all race-stratified analyses due to structural CDC suppression of race-specific hospitalization data. 95% confidence intervals derived by nonparametric bootstrap resampling of season-level pairs (1,000 iterations, percentile method). Cross-phase comparisons of absolute rate levels should be interpreted with caution due to a denominator discontinuity between bridged (PRE) and unbridged (DISR, REC) population estimates; within-phase rate ratios are internally consistent. CI = confidence interval; AIAN = American Indian/Alaska Native; API = Asian/Pacific Islander.

**Figure 3.**
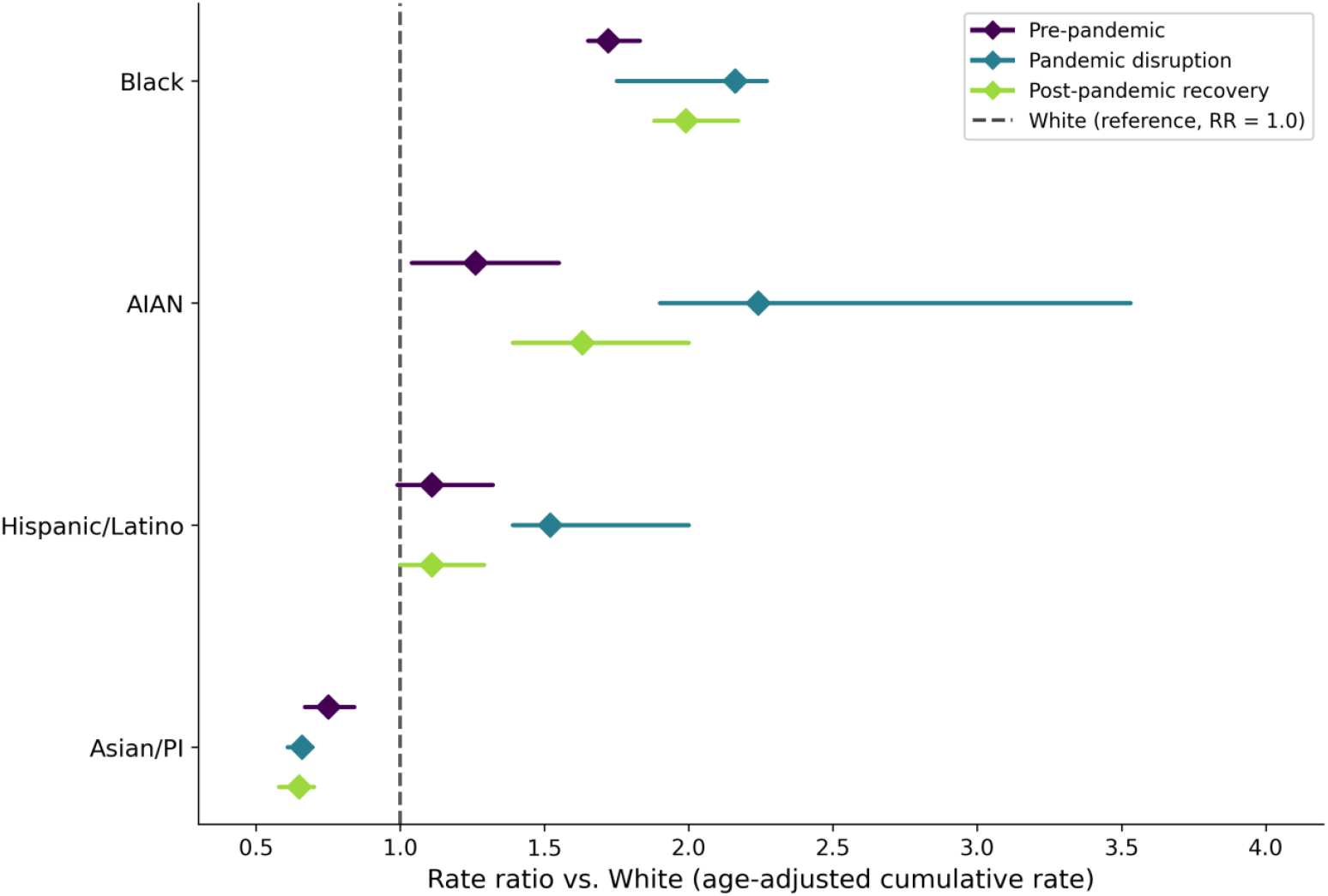
Age-adjusted influenza hospitalization rate ratios by race/ethnicity and epidemiological phase, FluSurv-NET surveillance network, United States, 2009–2025.

### 3.3. Phase-level differences in mean weekly hospitalization rates

Pairwise effect sizes indicated large differences between the recovery and both prior phases: Cohen’s d was 1.11 for pre-pandemic versus recovery and 0.97 for disruption versus recovery, with a small effect between pre-pandemic and disruption (d = 0.17), consistent with the descriptive similarity between those phases.

Season-level OLS regression estimated a pre-pandemic intercept of 1.51 per 100,000 (95% CI 0.80–2.22, p < 0.001). Recovery-phase coefficients were positive across both model specifications (primary: β = 1.13, 95% CI −0.35 to 2.60, p = 0.12; sensitivity: β = 2.76, 95% CI −0.85 to 6.37, p = 0.12) but did not reach statistical significance, reflecting substantial between-season variability at this sample size. The gap between unadjusted and adjusted R^2^ in the primary model (0.203 vs. 0.071) is consistent with limited degrees of freedom at n = 15 seasons rather than model misspecification, and effect size estimation is foregrounded accordingly. The disruption-phase coefficient was near zero and non-significant in both specifications (primary: β = −0.16, p = 0.84). The sensitivity model, which adjusted for population denominator version and the 2009–10 pandemic season, produced directionally consistent estimates with reduced fit relative to the primary specification (adjusted R^2^ 0.057 vs. 0.071; SM3).

A linear mixed-effects model with a season-level random intercept yielded a random effect variance of zero (ICC = 0), confirming that phase classification determined entirely at the season level, which absorbed all between-season variance and precluded estimation of phase fixed effects. The within-season week coefficient was −0.055 per 100,000 per week (95% CI −0.074 to −0.037, p < 0.001), reflecting a consistent within-season decline from peak toward the inter-seasonal baseline (SM3).

### 3.4. Seasonal decomposition

STL decomposition revealed a marked structural shift in the seasonal character of influenza hospitalization between the pre-pandemic and post-pandemic recovery periods (Figure 4). In the pre-pandemic series (300 weeks; ten seasons), seasonality was weak (Fs = 0.137) and the dominant source of variation was unstructured residual noise (SD 1.64 per 100,000), with a moderate upward trend (Ft = 0.237; trend range 3.18 per 100,000) and a seasonal amplitude IQR of 1.23 per 100,000 (Figure 4A). In the post-pandemic recovery series (112 weeks; three seasons), this structure was substantially reorganized: strength of seasonality increased to 0.979, residual SD collapsed to 0.45 per 100,000, and the trend component was effectively absent (Ft = 0.003; trend range 0.15 per 100,000), with seasonal amplitude nearly tripling to 3.44 per 100,000 (Figure 4B); these estimates should be interpreted with caution given the limited series length, though the magnitude of the shift across all four decomposition metrics is consistent with a genuine structural change rather than an artifact of the shorter window. The recovery phase is thus characterized by a highly structured, repeating annual cycle with minimal residual noise and no discernible secular trend, in contrast to the heterogeneous, trend-driven pre-pandemic pattern.

**Figure 4.**
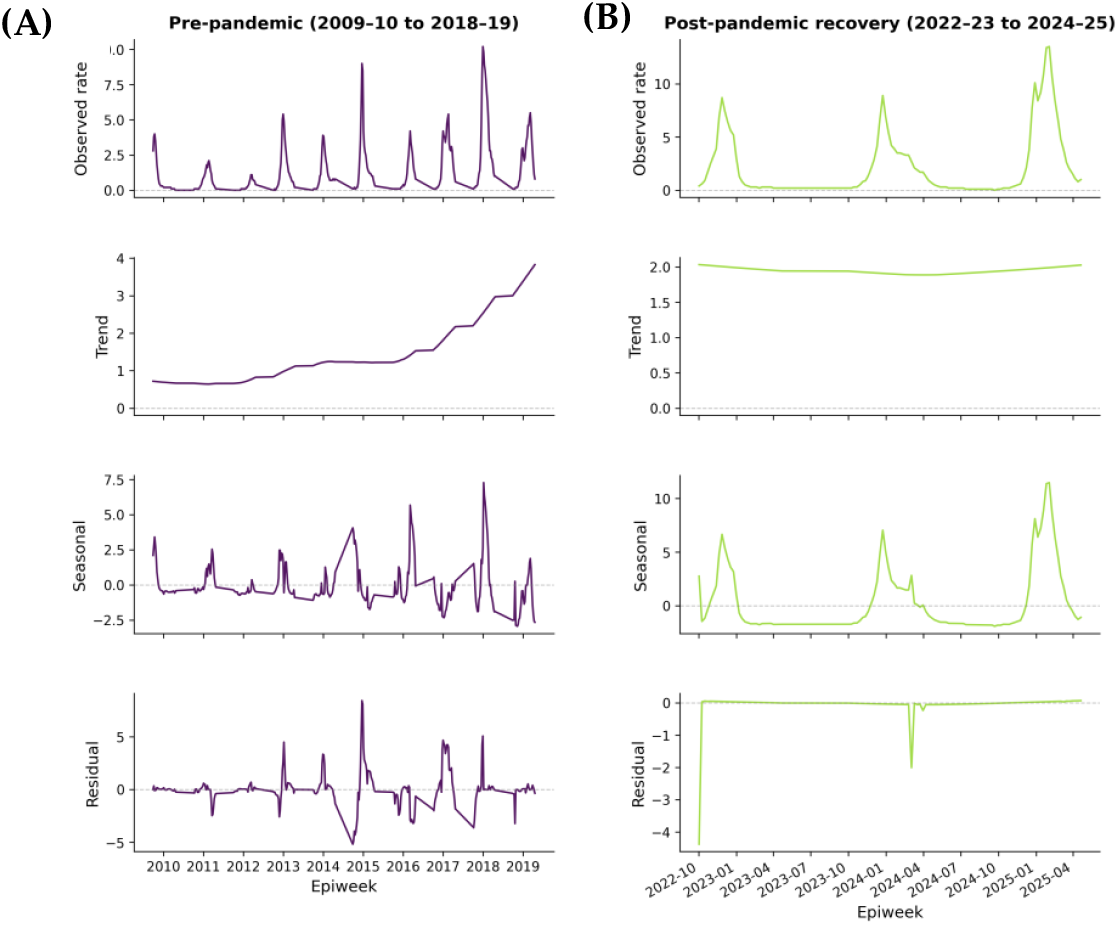
STL decomposition of weekly influenza hospitalization rates, FluSurv-NET, United States. Panel A: pre-pandemic series (2009–10 to 2018–19; n = 300 weeks). Panel B: post-pandemic recovery series (2022–23 to 2024–25; n = 112 weeks). Each panel shows the observed rate, extracted trend, seasonal, and residual components from top to bottom. Decomposition performed using robust LOESS with a 52-week period. All rates per 100,000 population. The pandemic disruption phase (2019–20 to 2021–22) was excluded from both series.

### 3.5. Time-series forecasting

A Prophet model trained on 300 weeks of pre-pandemic surveillance data (2009–10 to 2018–19) was used to generate a counterfactual baseline representing expected hospitalization rates absent pandemic disruption (Fig. 5). The primary specification used a flat growth assumption, holding the pre-pandemic rate level constant without trend extrapolation. Under this model, the first exceedance of the 95% prediction interval upper bound occurred in early February 2020, coinciding with the onset of pandemic-period disruption. The February 2020 detection date was identical across 9 of 10 leave-one-out refits excluding individual training seasons, with the sole exception advancing detection by 2 weeks, confirming the finding is not attributable to any single pre-pandemic season (SM4). At the season level, mean observed rates in the recovery phase exceeded the flat model projection across all three complete recovery seasons, with the largest mean gap in 2024–25 (observed 4.21 vs. projected 1.53 per 100,000; gap +2.68) and more modest exceedances in 2022–23 and 2023–24 (gaps +0.56 and +0.54 per 100,000, respectively). The disruption-phase gap was mixed: 2019–20 showed a modest positive mean gap (+0.67 per 100,000), while 2021–22 showed a large positive gap (+1.35 per 100,000), reflecting a projected baseline held at pre-pandemic levels against near-zero observed rates during the pandemic-associated collapse of influenza activity that season (SM4).

**Figure 5.**
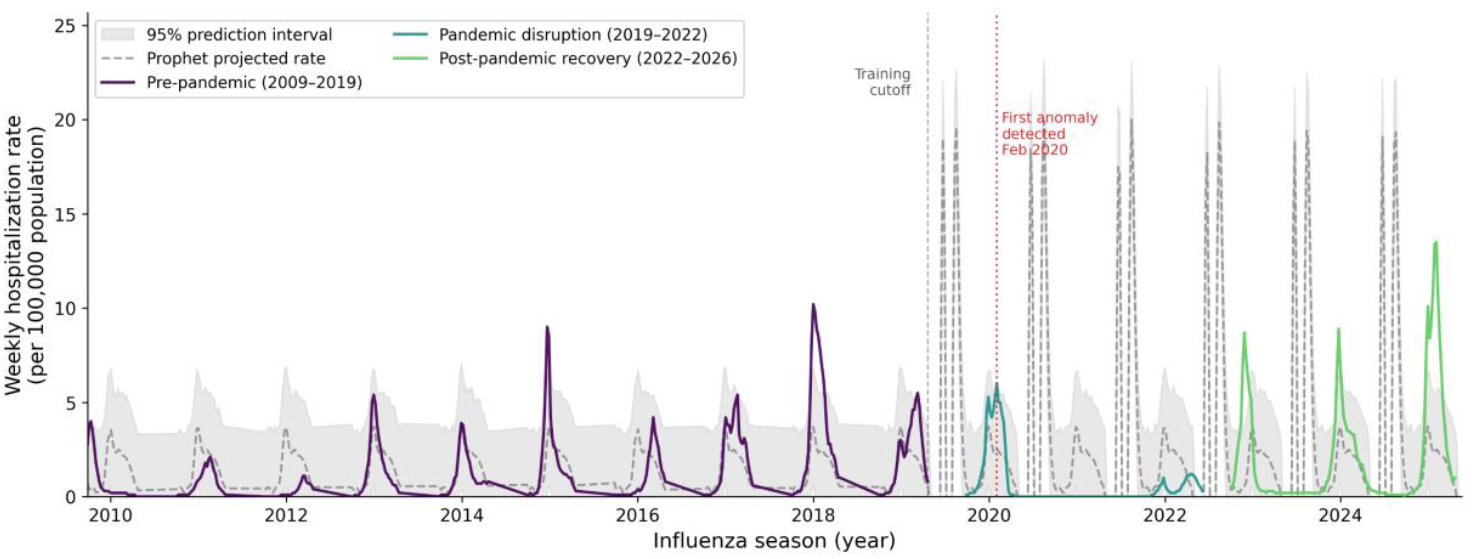
Prophet time-series forecast of weekly influenza hospitalization rates, FluSurv-NET, United States, 2009–2025. The primary flat growth model was trained on pre-pandemic surveillance data (2009–10 to 2018–19; n = 300 weeks). The dashed line represents the model-projected rate; shaded band represents the 95% prediction interval. Observed rates are shown by epidemiological phase. The vertical dotted line marks the first week in which observed rates exceeded the upper prediction bound (February 2020). The vertical dashed line marks the training cutoff (April 2019). All rates per 100,000 population.

**Figure 6.**
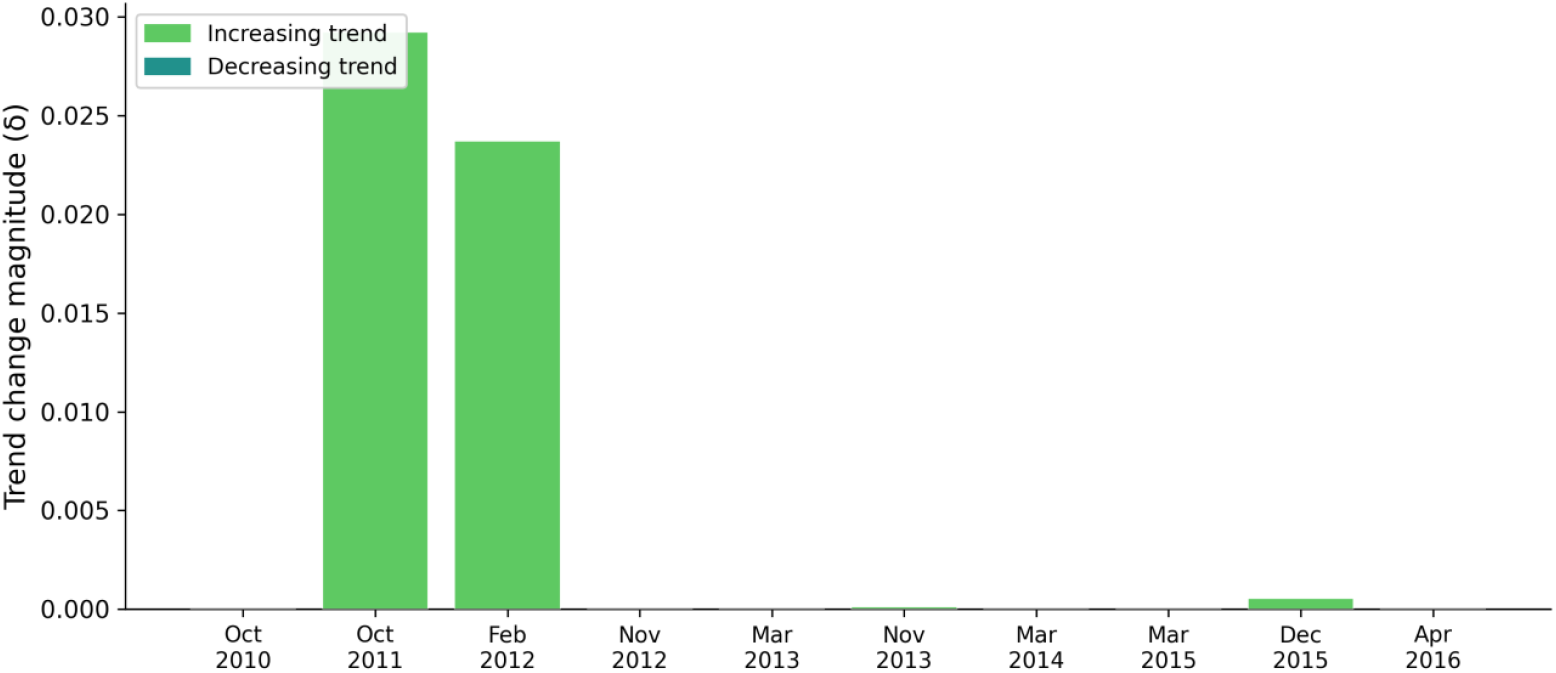
Prophet forecast sensitivity analysis: flat growth (primary) versus linear growth (sensitivity), FluSurv-NET, United States, 2009–2025. Both models trained on the pre-pandemic series (2009– 10 to 2018–19). Vertical dotted lines indicate first exceedance of the 95% upper prediction bound for each model (February 2020 and November 2022, respectively). The 146-week divergence in first detection date reflects systematic over-projection by the linear model in the post-pandemic period. All rates per 100,000 population. See SM4 for held-out validation metrics and season-level forecast gaps.

A sensitivity model using linear growth extrapolated the pre-pandemic upward trend into the forecast period, substantially elevating the projected baseline over time. Under this specification, first exceedance was delayed to November 2022, 146 weeks later than the flat model and well exceeding the pre-specified four-week convergence threshold. The linear model systematically over-projected mean rates in all recovery seasons (projected means 3.53 to 4.37 per 100,000 vs. observed 1.61 to 4.21 per 100,000), attributing observed burden to continued trend rather than anomalous elevation. Peak-level gap comparisons under the linear model are not reported for 2023–24, as the projected peak for that season reflects a forecasting artifact driven by its atypical 52-week duration rather than a meaningful model estimate. Given the flat model’s substantially superior held-out predictive performance on the 2018–19 validation season (MAE 1.01 vs. 1.59; R^2^ 0.23 vs. 0.005), the linear model’s delayed detection reflects over-projection from trend extrapolation rather than a later true onset of anomalous burden. Model growth specification sensitivity outlined in SM4.

### 3.6. Isolation Forest anomaly detection

The 2020–2021 season was excluded from Isolation Forest scoring due to CDC structural suppression of weekly hospitalization data during that period. Of the remaining 15 seasons, three were classified as robust anomalies, defined as flagged at all three contamination thresholds in the sensitivity sweep (0.05, 0.10, and 0.15): 2017–2018, 2023– 2024, and 2024–2025 (Fig. 7; SM4). The model was trained on pre-pandemic seasons (2009–10 to 2018–19) using six standardized season-level features: peak rate, mean rate, cumulative rate, season duration, peak week, and rate coefficient of variation.

**Figure 7.**
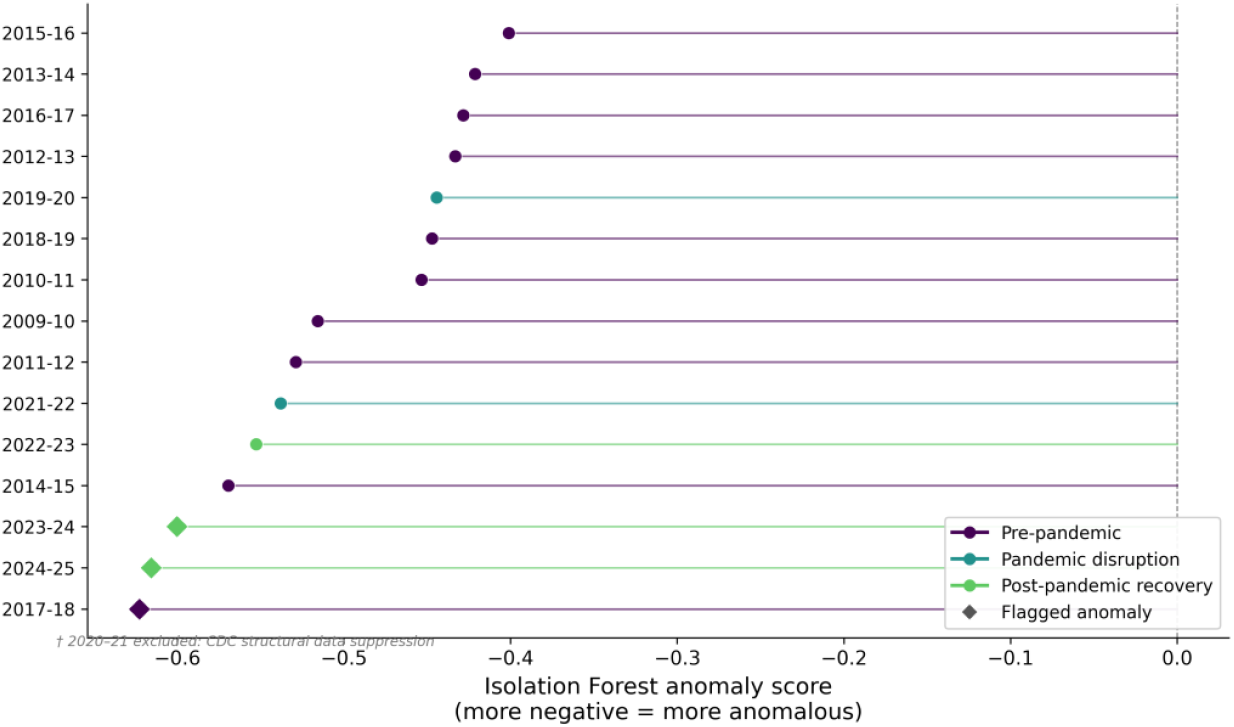
Isolation Forest anomaly scores by season, FluSurv-NET, United States, 2009–2025. Seasons are ranked by anomaly score from most to least anomalous (top to bottom). More negative scores indicate greater departure from the pre-pandemic normative distribution. Diamond markers indicate seasons classified as anomalous at the primary contamination threshold (0.10); circle markers indicate seasons classified as normal. Colors correspond to epidemiological phase. The 2020–21 season was excluded from scoring due to CDC structural suppression of weekly hospitalization data. Model trained on pre-pandemic seasons (2009–10 to 2018–19) with standardized features: peak rate, mean rate, cumulative rate, season duration, peak week, and rate coefficient of variation. See SM4 for full feature matrix and contamination sensitivity sweep.

The 2017–2018 pre-pandemic season received the most anomalous score in the dataset (−0.623), distinguished by the highest pre-pandemic peak rate (10.2 per 100,000), mean weekly rate (3.42 per 100,000), and cumulative rate (102.7 per 100,000) in the training distribution, with peak burden concentrated at MMWR week 1. The 2024–2025 and 2023–2024 seasons ranked second and third in anomaly score (−0.615 and −0.600, respectively), with peak rates of 13.5 and 8.9 per 100,000 and cumulative rates of 126.3 and 83.7 per 100,000, each exceeding the pre-pandemic normative distribution across multiple feature dimensions. The 2014–2015 season was flagged only at the highest contamination threshold (0.15) and is not considered a robust anomaly. All three robust anomalies showed concordant classification across both methods, with positive Prophet forecast gaps and Isolation Forest anomaly flags in each case, providing cross-method validation of the primary finding (SM4).

## 4. Discussion

Post-pandemic influenza in the United States does not represent a simple return to pre-pandemic norms. The recovery period captured through FluSurv-NET surveillance is instead characterized by compressed but substantially intensified seasonality, elevated hospitalization burden across demographic groups, and persistent racial and ethnic disparities that widened during the pandemic disruption period and have not fully resolved. These findings, derived retrospectively from a decade of population-based surveillance data using time-series decomposition, counterfactual forecasting, and unsupervised anomaly detection, are mutually reinforcing and collectively suggest that the structural character of influenza seasonality in the United States has undergone a meaningful reorganization in the post-pandemic period.

The burden elevation observed in the recovery phase is consistent with the immunological debt hypothesis, which holds that suppression of influenza transmission during the pandemic period through non-pharmaceutical interventions reduced opportunities for natural immune boosting at the population level, producing an expanded susceptible cohort when community mixing resumed [4–6]. Adults aged 65 and older carried the highest absolute burden across all three phases, and the post-pandemic amplification of that burden is epidemiologically consequential given the well-established relationship between influenza severity and age-related immune senescence [23–25]. The dramatic advance in median peak timing from late winter to early winter across all age groups and virus types adds a planning dimension to the burden finding: vaccination program schedules and healthcare surge frameworks calibrated to historical late-season peaks may be poorly aligned with the current seasonal pattern, and prospective monitoring is warranted to determine whether early-winter onset is a durable feature of the post-pandemic landscape or a transitional phenomenon [7,9,26].

The STL decomposition results provide quantitative support for a structural change in seasonal influenza dynamics that extends beyond burden elevation alone. Pre-pandemic influenza activity was dominated by residual noise, with weak and irregular seasonality consistent with heterogeneous epidemic dynamics driven by shifting antigenic profiles and variable population immunity across seasons. The post-pandemic recovery period presents a qualitatively different picture: near-perfect seasonal regularity, negligible residual variance, and no discernible secular trend. This transition from a noise-dominated to a seasonality-dominated signal is epidemiologically interpretable. Pandemic-period suppression of influenza circulation likely produced a susceptible population with reduced prior-season immunity heterogeneity, enabling more uniform and temporally predictable seasonal dynamics upon re-emergence [27–29]. From a preparedness standpoint, higher-amplitude but more predictable annual cycles could support more precise surge planning and resource allocation if the pattern persists. Whether this regularity reflects a durable post-pandemic immune landscape shift or a transitional period remains an open question; the recovery-phase observation window spans only three complete seasons, and between-season variability in vaccine composition, uptake, and antigenic match will continue to influence the degree of structural regularity observed in future seasons.

The racial and ethnic disparity findings extend a well-documented pattern of inequitable influenza burden with new post-pandemic context [10,11,30]. Black persons experienced the highest and most consistently elevated hospitalization rates relative to the White reference group across all three phases. The disparity widened during the disruption period and attenuated only partially in recovery, consistent with longstanding racial disparities in infectious disease hospitalization burden documented across seasons in the United States [10,31]. American Indian and Alaska Native (AI/AN) persons showed the largest disruption-phase elevation of any group examined, though this estimate carries substantial uncertainty reflecting the limited number of contributing seasons. This pattern is consistent with documented disparities in infectious disease outcomes among Indigenous populations in the United States [32,33] and internationally, reflecting systemic rather than incidental inequities in burden distribution [34]. The disruption-phase widening for these groups maps onto structural conditions that became acutely visible during the COVID-19 period: populations with constrained ability to reduce occupational exposure, access timely care, or benefit consistently from vaccination programs faced compounded infectious disease burden when influenza co-circulated with SARS-CoV-2 [35,36]. The partial return of Hispanic and Latino rates toward pre-pandemic levels in recovery, in contrast to the persistence of elevated Black and AI/AN rates, suggests heterogeneity in disparity mechanisms and trajectories across groups that aggregate rate ratios cannot fully characterize. The present analysis is descriptive and does not adjust for vaccination status, comorbidity burden, or healthcare access; the rate ratios reported here reflect population-level burden distribution rather than etiologic attribution.

The convergent findings from Prophet forecasting and Isolation Forest anomaly detection strengthen the inferential case for recovery-phase burden elevation beyond what descriptive statistics alone could support. The two methods approach the problem differently: Prophet evaluates week-level exceedance of a pre-pandemic prediction interval, while Isolation Forest scores season-level feature vectors against a pre-pandemic normative distribution. Both independently identified the 2023-2024 and 2024-2025 seasons as anomalous. The consistent flagging of the 2017-2018 season as the sole pre-pandemic anomaly by both methods provides internal validation, confirming sensitivity to genuine epidemiological outliers rather than recovery-phase artifacts; that season was previously characterized in the literature as unusually severe and was associated with predominant A(H3N2) circulation [37]. The 146-week divergence in first detection date between flat and linear growth Prophet specifications illustrates the degree to which surveillance-oriented forecasting is sensitive to baseline trend assumptions, and underscores the importance of prospective model validation before deployment in operational settings.

Several limitations of this analysis warrant acknowledgment. FluSurv-NET covers approximately 10% of the US population across 14 states and is not designed to produce nationally representative rate estimates; regional heterogeneity in influenza burden remains uncharacterized. The season-level OLS regression is based on 15 seasons and is underpowered to detect modest phase differences; effect size estimates are foregrounded over p-values accordingly. The disparity analysis presents descriptive age-adjusted rate ratios and does not adjust for vaccination status, comorbidity prevalence, or healthcare access, and should not be interpreted as an etiologic analysis of disparity mechanisms. The population denominator transition from bridged to unbridged race and ethnicity estimates at the 2020-2021 boundary limits direct cross-phase comparison of absolute rates, though within-phase rate ratios are internally consistent. The Isolation Forest model was trained on ten pre-pandemic seasons, and the small training set limits threshold precision; contamination sensitivity analyses were conducted to address this constraint, and findings were restricted to seasons flagged robustly across all levels. Subgroup-level anomaly detection stratified by age or race and ethnicity was outside the scope of the present analysis and represents a direction for future work.

## 5. Conclusions

Sixteen seasons of FluSurv-NET surveillance data demonstrate a post-pandemic influenza recovery characterized by substantially elevated hospitalization burden, a structural shift toward compressed and highly regular annual seasonality, earlier-onset seasonal peaks, and anomalous severity in the two most recent complete seasons identified independently by time-series forecasting and unsupervised anomaly detection. Racial and ethnic disparities in hospitalization rates were substantial prior to the pandemic, widened during the disruption period, and have not returned to pre-pandemic levels for several groups, reflecting heterogeneity in underlying population risk that aggregate burden estimates alone do not capture. Whether the elevated and reorganized recovery-phase pattern represents a transient resurgence or a more durable shift in the influenza epidemiological baseline cannot be determined from three recovery seasons alone. The analytic approach applied here, combining decomposition, counterfactual forecasting, and anomaly detection against a pre-pandemic normative baseline, is applicable to other population-based surveillance systems navigating similar post-pandemic questions. Continued surveillance with standardized methodology across future seasons remains essential for characterizing the trajectory of influenza burden and informing preparedness planning in the post-pandemic era.

## Data Availability

All data analyzed in this study are publicly available. Influenza hospitalization surveillance data were obtained from the CDC FluSurv-NET Influenza Hospitalization Surveillance Network and are accessible through the CDC FluView Interactive platform at https://www.cdc.gov/fluview/overview/influenza-hospitalization-surveillance.html.

https://github.com/h-hedman/healthcare-data-science/tree/main/fluserv-net-surveillance

## Supplementary Materials

SM1: Analysis code and reproducibility; GitHub repository link. SM2: Table S1: Season-level influenza hospitalization descriptive statistics, FluSurv-NET, 2009-2025. Figure S1: Season-level influenza hospitalization trajectories by virus type and epidemiological phase. SM3: Table S2a: OLS regression of mean weekly influenza hospitalization rate on epidemiological phase, primary and sensitivity models. Table S2b: Linear mixed-effects model within-season weekly rate trend. SM4: Table S3: Prophet held-out validation metrics, flat versus linear growth. Figure S2: Prophet sensitivity model comparison, linear versus flat growth. Table S4a: Isolation Forest season-level feature matrix and anomaly classifications. Table S4b: Isolation Forest contamination parameter sensitivity sweep. Figure S3: Prophet flat growth model held-out validation, 2018-19 season. Table S4c: Leave-one-out detection date stability, Prophet flat growth model. Table S4d: Cross-method convergence of Prophet forecast gaps and Isolation Forest anomaly classifications.

## Institutional Review Board Statement

Not applicable.

## Acknowledgments

During the preparation of this manuscript, the author used Claude (Anthropic, claude.ai) for assistance with manuscript drafting and editing. The author reviewed and edited all output and takes full responsibility for the content of this publication.

## Disclaimer/Publisher’s Note

The statements, opinions and data contained in all publications are solely those of the individual author(s) and contributor(s) and not of MDPI and/or the editor(s). MDPI and/or the editor(s) disclaim responsibility for any injury to people or property resulting from any ideas, methods, instructions or products referred to in the content.

## References

1. Hanage, W.P.; Schaffner, W. Burden of Acute Respiratory Infections Caused by Influenza Virus, Respiratory Syncytial Virus, and SARS-CoV-2 with Consideration of Older Adults: A Narrative Review. Infect Dis Ther 2025, 14, 5–37, doi:10.1007/s40121-024-01080-4.

2. Tanner, A.R.; Dorey, R.B.; Brendish, N.J.; Clark, T.W. Influenza Vaccination: Protecting the Most Vulnerable. European Respiratory Review 2021, 30, doi:10.1183/16000617.0258-2020.

3. CDC Influenza Hospitalization Surveillance Network (FluSurv-NET) Available online: https://www.cdc.gov/flu-view/overview/influenza-hospitalization-surveillance.html (accessed on 11 March 2026).

4. Wu, E.; Wu, V.; Wu, K.-H.; Wu, K.-C.; Huang, J.-Y. Immunity Debt Regarding the Aspect of Influenza in the Post-COVID-19 Era in Taiwan. Viruses 2024, 16, doi:10.3390/v16091468.

5. Leung, C.; Su, L. Postpandemic Immunity Debt of Common Cold in England: An Interrupted Time Series Study. International Journal of Infectious Diseases 2025, 156, 107918, doi:10.1016/j.ijid.2025.107918.

6. Leung, C.; Konya, L.; Su, L. Postpandemic Immunity Debt of Influenza in the USA and England: An Interrupted Time Series Study. Public Health 2024, 227, 239–242, doi:10.1016/j.puhe.2023.12.009.

7. Presti, S.; Manti, S.; Gambilonghi, F.; Parisi, G.F.; Papale, M.; Leonardi, S. Comparative Analysis of Pediatric Hospitalizations during Two Consecutive Influenza and Respiratory Virus Seasons Post-Pandemic. Viruses 2023, 15, doi:10.3390/v15091825.

8. Giovanetti, M.; Ali, S.; Slavov, S.N.; Azarian, T.; Cella, E. Epidemiological Transitions in Influenza Dynamics in the United States: Insights from Recent Pandemic Challenges. Microorganisms 2025, 13, doi:10.3390/microorganisms13030469.

9. Lee, S.S.; Viboud, C.; Petersen, E. Understanding the Rebound of Influenza in the Post COVID-19 Pandemic Period Holds Important Clues for Epidemiology and Control. International Journal of Infectious Diseases 2022, 122, 1002–1004, doi:10.1016/j.ijid.2022.08.002.

10. Lippert, J.F.; Buscemi, J.; Saiyed, N.; Silva, A.; Benjamins, M.R. Influenza and Pneumonia Mortality Across the 30 Biggest U.S. Cities: Assessment of Overall Trends and Racial Inequities. J. Racial and Ethnic Health Disparities 2022, 9, 1152–1160, doi:10.1007/s40615-021-01056-x.

11. Jones, E.A.K.; Mitra, A.K.; Malone, S. Racial Disparities and Common Respiratory Infectious Diseases in Children of the United States: A Systematic Review and Meta-Analysis. Diseases 2023, 11, doi:10.3390/diseases11010023.

12. Dawood, F.S.; Chaves, S.S.; Pérez, A.; Reingold, A.; Meek, J.; Farley, M.M.; Ryan, P.; Lynfield, R.; Morin, C.; Baumbach, J.; et al. Complications and Associated Bacterial Coinfections Among Children Hospitalized With Seasonal or Pandemic Influenza, United States, 2003–2010. J Infect Dis 2014, 209, 686–694, doi:10.1093/infdis/jit473.

13. Jeganathan, N.; Grewal, S.; Sathananthan, M. Comparison of Deaths from COVID-19 and Seasonal Influenza in the USA. Lung 2021, 199, 559–561, doi:10.1007/s00408-021-00468-0.

14. Solomon, D.A.; Sherman, A.C.; Kanjilal, S. Influenza in the COVID-19 Era. JAMA 2020, 324, 1342–1343, doi:10.1001/jama.2020.14661.

15. Meites, E.; Knuth, M.; Hall, K.; Dawson, P.; Wang, T.W.; Wright, M.; Yu, W.; Senesie, S.; Stephenson, E.; Imachukwu, C.; et al. COVID-19 Scientific Publications From the Centers for Disease Control and Prevention, January 2020–January 2022. Public Health Rep 2023, 138, 241–247, doi:10.1177/00333549221134130.

16. Pervaiz, F.; Pervaiz, M.; Rehman, N.A.; Saif, U. FluBreaks: Early Epidemic Detection from Google Flu Trends. Journal of Medical Internet Research 2012, 14, e2102, doi:10.2196/jmir.2102.

17. Rosenström, T.; Jokela, M.; Puttonen, S.; Hintsanen, M.; Pulkki-Råback, L.; Viikari, J.S.; Raitakari, O.T.; Keltikan-gas-Järvinen, L. Pairwise Measures of Causal Direction in the Epidemiology of Sleep Problems and Depression. PLOS ONE 2012, 7, e50841, doi:10.1371/journal.pone.0050841.

18. Cleveland, R.B.; Cleveland, W.S.; McRae, J.E.; Terpenning, I. STL: A Seasonal-Trend Decomposition. Journal of Official Statistics 1990, 6, 3–73.

19. Hyndman, R.; Athanasopoulos, G. Forecasting: Principles and Practice. 2021.

20. Gao, X.; Qi, X.; Wan, Y.; Xu, Z.; Xie, L.; Hu, H. Prediction of Influenza-like Cases in Chaoyang District of Beijing Based on Prophet Model. In Proceedings of the Proceedings of the 14th International Conference on Information Communication and Applications; Association for Computing Machinery: New York, NY, USA, January 24 2026; pp. 16–21.

21. Taylor, S.J.; Letham, B. Forecasting at Scale. The American Statistician 2018, 72, 37–45, doi:10.1080/00031305.2017.1380080.

22. Liu, F.T.; Ting, K.M.; Zhou, Z.-H. Isolation Forest. In Proceedings of the 2008 Eighth IEEE International Conference on Data Mining; December 2008; pp. 413–422.

23. Langer, J.; Welch, V.L.; Moran, M.M.; Cane, A.; Lopez, S.M.C.; Srivastava, A.; Enstone, A.L.; Sears, A.; Markus, K.J.; Heuser, M.; et al. High Clinical Burden of Influenza Disease in Adults Aged ≥ 65 Years: Can We Do Better? A Systematic Literature Review. Adv Ther 2023, 40, 1601–1627, doi:10.1007/s12325-023-02432-1.

24. Jefferson, T.; Rivetti, D.; Rivetti, A.; Rudin, M.; Pietrantonj, C.D.; Demicheli, V. Efficacy and Effectiveness of Influenza Vaccines in Elderly People: A Systematic Review. The Lancet 2005, 366, 1165–1174, doi:10.1016/S0140-6736(05)67339-4.

25. Antonelli Incalzi, R.; Consoli, A.; Lopalco, P.; Maggi, S.; Sesti, G.; Veronese, N.; Volpe, M. Influenza Vaccination for Elderly, Vulnerable and High-Risk Subjects: A Narrative Review and Expert Opinion. Intern Emerg Med 2024, 19, 619–640, doi:10.1007/s11739-023-03456-9.

26. Nisar, N.; Badar, N.; Safdar, I. Assessing Influenza Activity Variations in the Asian Region during the Pre-and Post-Pandemic Period (2017–2023). PLOS ONE 2025, 20, e0323465, doi:10.1371/journal.pone.0323465.

27. Nofzinger, T.B.; Huang, T.T.; Lingat, C.E.R.; Amonkar, G.M.; Edwards, E.E.; Yu, A.; Smith, A.D.; Gayed, N.; Gad-dey, H.L. Vaccine Fatigue and Influenza Vaccination Trends across Pre-, Peri-, and Post-COVID-19 Periods in the United States Using Epic’s Cosmos Database. PLOS ONE 2025, 20, e0326098, doi:10.1371/journal.pone.0326098.

28. Wang, Q.; Jia, M.; Jiang, M.; Cao, Y.; Dai, P.; Yang, J.; Yang, X.; Xu, Y.; Yang, W.; Feng, L. Increased Population Susceptibility to Seasonal Influenza during the COVID-19 Pandemic in China and the United States. Journal of Medical Virology 2023, 95, e29186, doi:10.1002/jmv.29186.

29. Gentile, A.; Juárez, M. del V.; Ensinck, G.; Lopez, O.; Melonari, P.; Fernández, T.; Gioiosa, A.; Lazarte, G.; Lobertti, S.; Lucion, M.F.; et al. Comparative Analysis of Influenza Epidemiology Before and After the COVID-19 Pandemic in Argentina (2018–2019 vs. 2022–2023). Influenza and Other Respiratory Viruses 2025, 19, e70078, doi:10.1111/irv.70078.

30. D’Adamo, A.; Schnake-Mahl, A.; Mullachery, P.H.; Lazo, M.; Diez Roux, A.V.; Bilal, U. Health Disparities in Past Influenza Pandemics: A Scoping Review of the Literature. SSM - Population Health 2023, 21, 101314, doi:10.1016/j.ssmph.2022.101314.

31. Adams, K.; Yousey-Hindes, K.; Bozio, C.H.; Jain, S.; Kirley, P.D.; Armistead, I.; Alden, N.B.; Openo, K.P.; Witt, L.S.; Monroe, M.L.; et al. Social Vulnerability, Intervention Utilization, and Outcomes in US Adults Hospitalized With Influenza. JAMA Netw Open 2024, 7, e2448003, doi:10.1001/jamanetworkopen.2024.48003.

32. Ehrenpreis, J.E.; Ehrenpreis, E.D. A Historical Perspective of Healthcare Disparity and Infectious Disease in the Native American Population. The American Journal of the Medical Sciences 2022, 363, 288–294, doi:10.1016/j.am-jms.2022.01.005.

33. Kruse, G.; Lopez-Carmen, V.A.; Jensen, A.; Hardie, L.; Sequist, T.D. The Indian Health Service and American Indian/Alaska Native Health Outcomes. Annual Review of Public Health 2022, 43, 559–576, doi:10.1146/annurev-publhealth-052620-103633.

34. Betts, J.M.; Weinman, A.L.; Oliver, J.; Braddick, M.; Huang, S.; Nguyen, M.; Miller, A.; Tong, S.Y.C.; Gibney, K.B. Influenza-Associated Hospitalisation and Mortality Rates among Global Indigenous Populations; a Systematic Review and Meta-Analysis. PLOS Global Public Health 2023, 3, e0001294, doi:10.1371/journal.pgph.0001294.

35. Golpour, M.; Jalali, H.; Alizadeh-Navaei, R.; Talarposhti, M.R.; Mousavi, T.; Ghara, A.A.N. Co-Infection of SARS-CoV-2 and Influenza A/B among Patients with COVID-19: A Systematic Review and Meta-Analysis. BMC Infect Dis 2025, 25, 145, doi:10.1186/s12879-025-10521-5.

36. Manno, M.; Pavia, G.; Gigliotti, S.; Pantanella, M.; Barreca, G.S.; Peronace, C.; Gallo, L.; Trimboli, F.; Colosimo, E.; Lamberti, A.G.; et al. Respiratory Virus Prevalence Across Pre-, During-, and Post-SARS-CoV-2 Pandemic Periods. Viruses 2025, 17, doi:10.3390/v17081040.

37. Potter, B.I.; Kondor, R.; Hadfield, J.; Huddleston, J.; Barnes, J.; Rowe, T.; Guo, L.; Xu, X.; Neher, R.A.; Bedford, T.; et al. Evolution and Rapid Spread of a Reassortant A(H3N2) Virus That Predominated the 2017–2018 Influenza Season. Virus Evol 2019, 5, vez046, doi:10.1093/ve/vez046.

