## Supplementary material for "Game Over for the Baseline: Anomalous Burden and Structural Seasonal Shifts in Post-Pandemic U.S. Influenza Hospitalization, 2009–2025": SM1-4

**Supplemental Materials 1 – Analysis Code and Reproducibility**

The code used for this study is available in the public repository at: https://github.com/h-hedman/healthcare-data-science/tree/main/fluserv-net-surveillance.

All scripts required to reproduce the models, tables, and figures are provided.

### Supplemental Materials 2 – Descriptive and Season-Level Detail

**Table S1**. Season-level influenza hospitalization descriptive statistics, FluSurv-NET: 2009–2025.

| Season | Phase | Denom | Partial | H1N1 Season | Weeks (n) | Active Weeks (n) | Median Rate | Q1 | Q3 | Mean Rate | SD | Peak Rate | Peak Week | EOS Cum Rate | EOS Age-Adj Rate |
|---|---|---|---|---|---|---|---|---|---|---|---|---|---|---|---|
| 2009-10 | PRE | bridged | 0 | 1 | 30 | 28 | 0.2 | 0.2 | 0.73 | 0.83 | 1.23 | 4 | 42 | 29.3 | 25.75 |
| 2010-11 | PRE | bridged | 0 | 0 | 30 | 29 | 0.45 | 0.1 | 1.2 | 0.72 | 0.67 | 2.1 | 8 | 21.5 | 19.76 |
| 2011-12 | PRE | bridged | 0 | 0 | 30 | 20 | 0.1 | 0 | 0.55 | 0.29 | 0.35 | 1.1 | 11 | 8.7 | 7.65 |
| 2012-13 | PRE | bridged | 0 | 0 | 30 | 27 | 0.85 | 0.22 | 1.98 | 1.47 | 1.6 | 5.4 | 1 | 44 | 40.86 |
| 2013-14 | PRE | bridged | 0 | 0 | 30 | 29 | 0.8 | 0.45 | 1.68 | 1.17 | 1.12 | 3.9 | 1 | 35.1 | 32.4 |
| 2014-15 | PRE | bridged | 0 | 0 | 30 | 30 | 1.2 | 0.35 | 2.45 | 1.93 | 2.32 | 9 | 52 | 64.1 | 55.86 |
| 2015-16 | PRE | bridged | 0 | 0 | 30 | 30 | 0.4 | 0.1 | 1.78 | 1.05 | 1.23 | 4.2 | 10 | 31.4 | 28.54 |
| 2016-17 | PRE | bridged | 0 | 0 | 30 | 30 | 1.8 | 0.32 | 3.38 | 2.07 | 1.75 | 5.4 | 8 | 62 | 56.16 |
| 2017-18 | PRE | bridged | 0 | 0 | 30 | 30 | 2.25 | 0.85 | 6.02 | 3.42 | 3.33 | 10.2 | 1 | 102.9 | 70.71 |
| 2018-19 | PRE | bridged | 0 | 0 | 30 | 30 | 2 | 0.38 | 3.38 | 2.12 | 1.75 | 5.5 | 11 | 63.6 | 59.99 |
| 2019-20 | DISR | bridged | 0 | 0 | 30 | 29 | 1.3 | 0.2 | 4.47 | 2.2 | 2.15 | 6 | 6 | 66 | 62.43 |
| 2020-21 | DISR | unbridged | 0 | 0 | 31 | 0 | | | | | | | 1 | 0.8 | 0.77 |
| 2021-22 | DISR | unbridged | 0 | 0 | 36 | 31 | 0.4 | 0.2 | 0.72 | 0.49 | 0.38 | 1.2 | 16 | 17.5 | 16.62 |
| 2022-23 | REC | unbridged | 0 | 0 | 30 | 30 | 0.55 | 0.3 | 3.15 | 2.08 | 2.6 | 8.7 | 48 | 62.4 | 59.13 |
| 2023-24 | REC | unbridged | 0 | 0 | 52 | 51 | 0.5 | 0.2 | 2.75 | 1.61 | 2.06 | 8.9 | 52 | 83.5 | 80.2 |

| | | | | | | | | | | | | | | | |
|---|---|---|---|---|---|---|---|---|---|---|---|---|---|---|---|
| 2024-25 | REC | unbridged | 0 | 0 | 30 | 30 | 2.05 | 0.52 | 8.22 | 4.21 | 4.43 | 13.5 | 6 | 126.2 | 121.95 |
| 2025-26 | REC | unbridged | 1 | 0 | 30 | 21 | 2.8 | 0.4 | 3.5 | 3.27 | 3.79 | 13.3 | 52 | 76 | 71.77 |

Season-level influenza hospitalization summary statistics for the overall FluSurv-NET surveillance network, 2009–2025. One row per season. All rates per 100,000 population. Phase abbreviations: PRE = pre-pandemic baseline (2009–10 to 2018–19); DISR = pandemic disruption (2019–20 to 2021–22); REC = post-pandemic recovery (2022–23 to 2024–25). Active weeks defined as weeks with a weekly hospitalization rate ≥0.1 per 100,000. The 2020–21 season median, mean, peak, and Q1/Q3 fields are blank due to structural CDC suppression of weekly hospitalization data; end-of-season cumulative rate is available for that season. Denom = population denominator version (bridged = pre-2020 bridged race estimates; unbridged = post-2020 unbridged estimates). Partial = partial season flag (1 = incomplete season at time of analysis). EOS = end of season. Q1/Q3 = 25th/75th percentile of weekly rates.

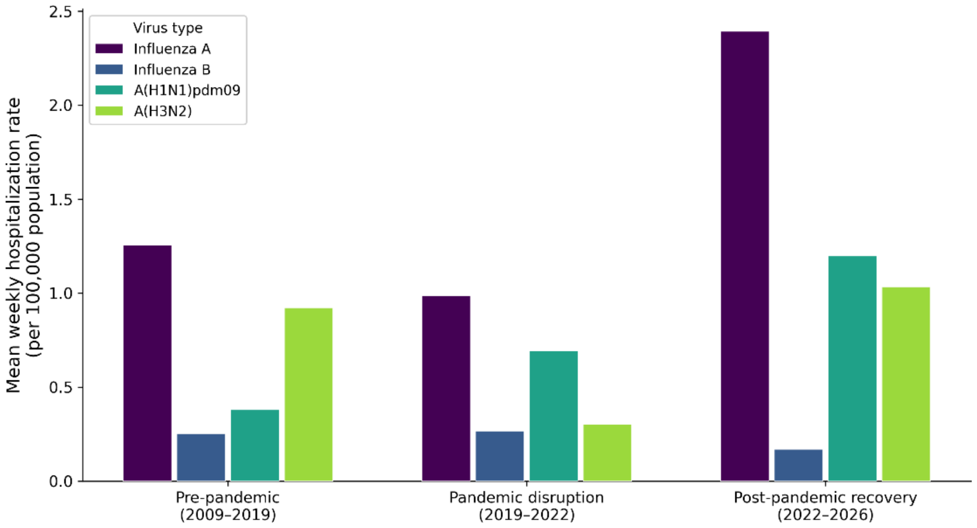


**Figure S1**. Season-level influenza hospitalization trajectories by virus type and epidemiological phase.

### Supplemental Materials 3 – Phase-Level Statistical Models

**Table S2a**. Ordinary least squares regression of mean weekly influenza hospitalization rate on epidemiological phase: primary and sensitivity models, FluSurv-NET, United States, 2009–2025.

| **Model** | **Parameter** | **Estimate** | **SE** | **95% CI Lower** | **95% CI Upper** | **t** | **p** | **$R^2$** | **Adj $R^2$** | **Seasons (n)** |
|---|---|---|---|---|---|---|---|---|---|---|
| Primary | Intercept (pre-pandemic reference) | 1.507 | 0.325 | 0.798 | 2.216 | 4.631 | 0.0006 | 0.203 | 0.071 | 15 |
| Primary | Pandemic disruption vs pre-pandemic (DISR) | -0.164 | 0.797 | -1.901 | 1.573 | -0.206 | 0.8402 | 0.203 | 0.071 | 15 |
| Primary | Post-pandemic recovery vs pre-pandemic (REC) | 1.127 | 0.677 | -0.349 | 2.603 | 1.664 | 0.122 | 0.203 | 0.071 | 15 |
| Sensitivity | Intercept (pre-pandemic reference) | 1.582 | 0.346 | 0.812 | 2.352 | 4.578 | 0.001 | 0.326 | 0.057 | 15 |
| Sensitivity | Pandemic disruption vs pre-pandemic (DISR) | 0.615 | 1.093 | -1.82 | 3.05 | 0.563 | 0.5861 | 0.326 | 0.057 | 15 |
| Sensitivity | Post-pandemic recovery vs pre-pandemic (REC) | 2.76 | 1.621 | -0.851 | 6.372 | 1.703 | 0.1194 | 0.326 | 0.057 | 15 |
| Sensitivity | Denominator version (unbridged vs bridged) | -1.708 | 1.466 | -4.974 | 1.559 | -1.165 | 0.2711 | 0.326 | 0.057 | 15 |
| Sensitivity | 2009–10 H1N1 pandemic season indicator | -0.749 | 1.093 | -3.183 | 1.686 | -0.685 | 0.5089 | 0.326 | 0.057 | 15 |

**Table S2b**. Linear mixed-effects model fixed effects within-season weekly influenza hospitalization rate trend, FluSurv-NET, United States, 2009–2025.

| Parameter | Estimate | SE | 95% CI Lower | 95% CI Upper | Seasons (n) |
|---|---|---|---|---|---|
| Intercept (pre-pandemic reference) | 0 | 10820536.78 | -21207862.38 | 21207862.38 | 15 |
| Within-season week number (linear trend) | -0.055 | 0.009 | -0.074 | -0.037 | 15 |

Intercept confidence interval reflects degenerate random-effect variance (ICC = 0); the fixed intercept is not interpretable.

### Supplemental Materials 4 – Time-Series Forecasting and Anomaly Detection

**Table S3**. Prophet held-out validation metrics: flat vs linear growth specification

| Model | MAR | RMSE | MAPE (%) | $R^2$ | Test Weeks (n) |
|---|---|---|---|---|---|
| Prophet_linear | 1.5897 | 1.7158 | — | 0.0053 | 30 |
| Prophet_flat | 1.0064 | 1.5079 | 86.14 | 0.2317 | 30 |

MAPE not reported for the linear growth model; unstable due to trend extrapolation over near-zero inter-seasonal observations.

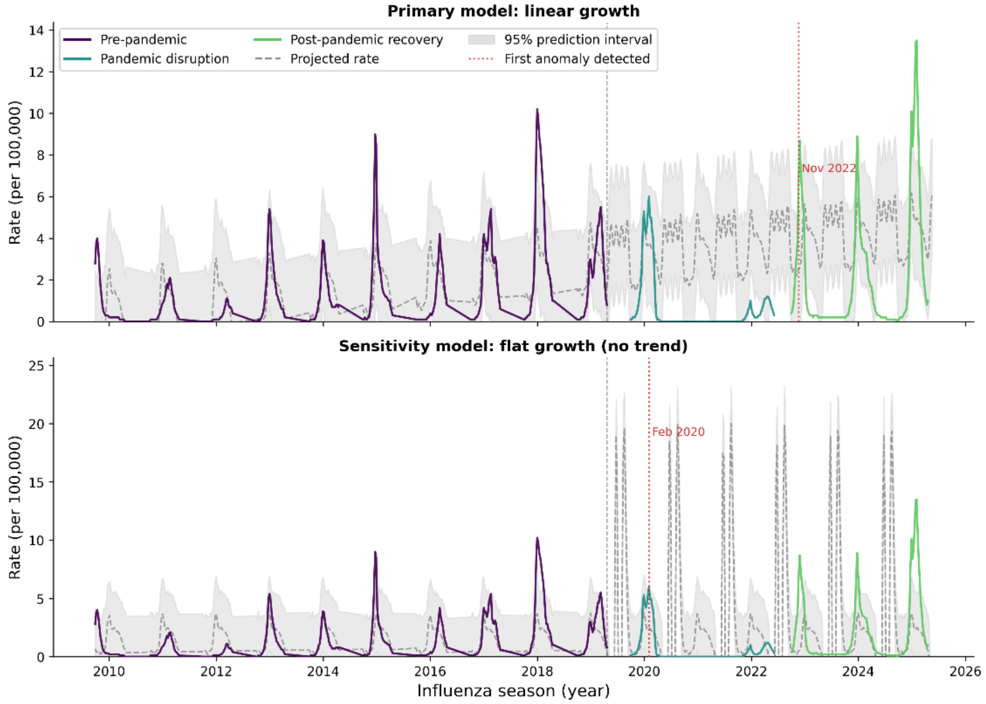


**Figure S2**. Prophet sensitivity model comparison: linear vs flat growth trend specification

### Supplemental Materials 4 – Isolation Forest Sensitivity Analyses

The Isolation Forest model was trained on pre-pandemic seasons (2009–10 to 2018–19) to establish a normative burden distribution, with scoring applied to all remaining seasons. The 2020–21 season was excluded from both training and scoring; CDC-suppressed near-zero rates during that period reflect a reporting artifact rather than true disease burden, and inclusion would produce a misleadingly low anomaly score.

**Table S4a**. Isolation Forest season-level feature matrix and anomaly classifications, FluSurv-NET, United States, 2009–2025 (2020–21 excluded).

| Season | Phase | Rank | Classification | Anomaly Score | Peak Rate | Mean Rate | Cum Rate | Weeks (n) | Peak Week | Rate CV |
|---|---|---|---|---|---|---|---|---|---|---|
| 2017-18 | PRE | 1 | Anomaly | -0.623 | 10.2 | 3.423 | 102.7 | 30 | 1 | 0.972 |
| 2024-25 | REC | 2 | Anomaly | -0.615 | 13.5 | 4.21 | 126.3 | 30 | 6 | 1.051 |
| 2023-24 | REC | 3 | Anomaly | -0.6 | 8.9 | 1.61 | 83.7 | 52 | 52 | 1.277 |
| 2014-15 | PRE | 4 | Normal | -0.569 | 9 | 1.93 | 57.9 | 30 | 30 | 1.2 |
| 2022-23 | REC | 5 | Normal | -0.553 | 8.7 | 2.083 | 62.5 | 30 | 26 | 1.247 |
| 2021-22 | DISR | 6 | Normal | -0.538 | 1.2 | 0.489 | 17.6 | 36 | 16 | 0.787 |
| 2011-12 | PRE | 7 | Normal | -0.529 | 1.1 | 0.287 | 8.6 | 30 | 11 | 1.211 |
| 2009-10 | PRE | 8 | Normal | -0.516 | 4 | 0.833 | 25 | 30 | 20 | 1.473 |
| 2010-11 | PRE | 9 | Normal | -0.453 | 2.1 | 0.723 | 21.7 | 30 | 8 | 0.928 |
| 2018-19 | PRE | 10 | Normal | -0.447 | 5.5 | 2.117 | 63.5 | 30 | 11 | 0.827 |
| 2019-20 | DISR | 11 | Normal | -0.444 | 6 | 2.197 | 65.9 | 30 | 6 | 0.977 |
| 2012-13 | PRE | 12 | Normal | -0.433 | 5.4 | 1.467 | 44 | 30 | 1 | 1.089 |
| 2016-17 | PRE | 13 | Normal | -0.428 | 5.4 | 2.067 | 62 | 30 | 8 | 0.845 |
| 2013-14 | PRE | 14 | Normal | -0.421 | 3.9 | 1.173 | 35.2 | 30 | 1 | 0.952 |
| 2015-16 | PRE | 15 | Normal | -0.401 | 4.2 | 1.05 | 31.5 | 30 | 10 | 1.169 |

The projected peak rate for 2023–24 under the linear growth model (19.3 per 100,000) reflects a forecasting artifact driven by that season's atypical 52-week reporting duration, which caused the model to extrapolate trend over an extended forecast horizon. This value is retained in the table for completeness but is not interpreted as a meaningful model estimate. The flat model projected peak for 2023–24 (3.23 per 100,000) is not affected by this artifact as it does not extrapolate trend.

**Table S4b**. Isolation Forest contamination parameter sensitivity sweep across three threshold levels, FluSurv-NET, United States, 2009–2025.

| Season | Phase | Primary Classification | Robust Anomaly | Primary Score | Contam=0.05 | Contam=0.10 | Contam=0.15 |
|---|---|---|---|---|---|---|---|
| 2017-18 | PRE | Anomaly | Yes | -0.623 | Anomaly | Anomaly | Anomaly |
| 2024-25 | REC | Anomaly | Yes | -0.615 | Anomaly | Anomaly | Anomaly |
| 2023-24 | REC | Anomaly | Yes | -0.6 | Anomaly | Anomaly | Anomaly |
| 2014-15 | PRE | Normal | No | -0.569 | Normal | Normal | Anomaly |
| 2022-23 | REC | Normal | No | -0.553 | Normal | Normal | Normal |
| 2021-22 | DISR | Normal | No | -0.538 | Normal | Normal | Normal |
| 2011-12 | PRE | Normal | No | -0.529 | Normal | Normal | Normal |
| 2009-10 | PRE | Normal | No | -0.516 | Normal | Normal | Normal |
| 2010-11 | PRE | Normal | No | -0.453 | Normal | Normal | Normal |
| 2018-19 | PRE | Normal | No | -0.447 | Normal | Normal | Normal |
| 2019-20 | DISR | Normal | No | -0.444 | Normal | Normal | Normal |
| 2012-13 | PRE | Normal | No | -0.433 | Normal | Normal | Normal |
| 2016-17 | PRE | Normal | No | -0.428 | Normal | Normal | Normal |
| 2013-14 | PRE | Normal | No | -0.421 | Normal | Normal | Normal |
| 2015-16 | PRE | Normal | No | -0.401 | Normal | Normal | Normal |
| 2020-21 | DISR | EXCLUDED | N/A — CDC data suppression | | EXCLUDED | EXCLUDED | EXCLUDED |

All changepoint delta values are near-zero by design. The flat growth specification contains no trend component; candidate changepoints therefore reflect seasonal timing adjustments only and do not indicate discrete rate-level shifts in the pre-pandemic series. The absence of large-magnitude changepoints confirms that no structural breaks in rate level were present in the training data under this specification, which is the expected and correct result for a flat growth model.

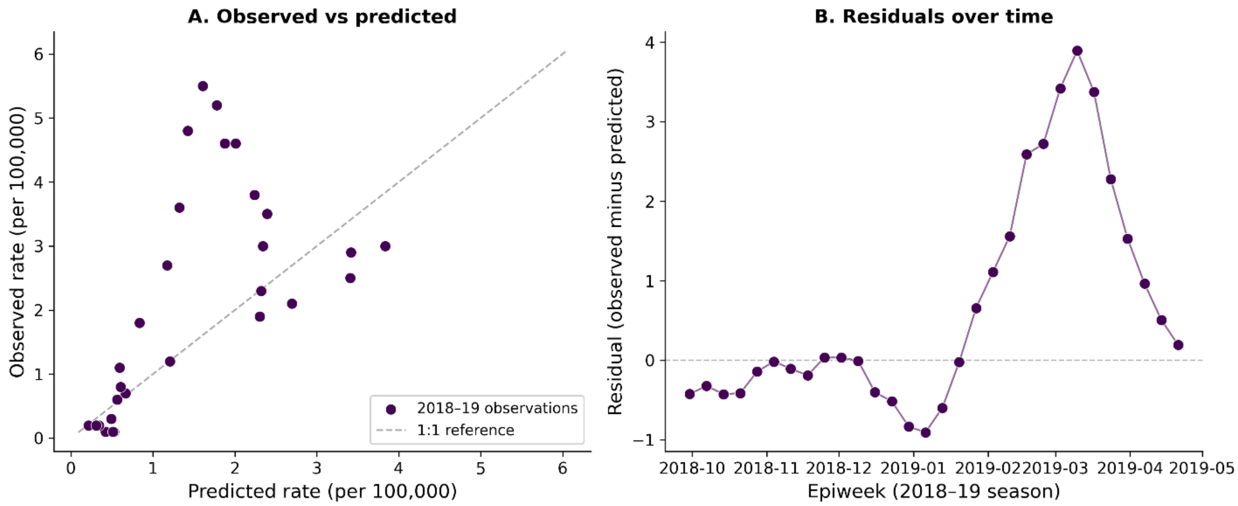


**Figure S3**. Held-out validation of the primary Prophet flat growth model, 2018–19 season, FluSurv-NET, United States. Panel A: observed versus predicted weekly hospitalization rates (per 100,000 population). The dashed line indicates perfect agreement (1:1 reference). Points above the reference line during peak weeks reflect the above-average severity of the 2018–19 season relative to the training baseline, consistent with conservative underprediction rather than model misspecification. Panel B: residuals (observed minus predicted) over time. The seasonal pattern of residuals, with negative values during the inter-seasonal period and positive values at peak, reflects expected within-season rate variation and shows no evidence of systematic temporal bias. Model trained on 2009–10 to 2017–18 (n = 270 weeks; n = 30 test weeks).

**Table S4b**. Leave-one-out detection date stability, Prophet flat growth model, FluSurv-NET, United States, 2009–2025.

| Excluded Season | Training Seasons (n) | First Detection Date | Gap vs Full Model (weeks) | Direction |
|---|---|---|---|---|
| None (full model) | 10 | 2/2/2020 | 0 | Reference |
| 2009-10 | 9 | 2/2/2020 | 0 | Same |
| 2010-11 | 9 | 2/2/2020 | 0 | Same |
| 2011-12 | 9 | 2/2/2020 | 0 | Same |
| 2012-13 | 9 | 2/2/2020 | 0 | Same |
| 2013-14 | 9 | 2/2/2020 | 0 | Same |
| 2014-15 | 9 | 2/2/2020 | 0 | Same |
| 2015-16 | 9 | 2/2/2020 | 0 | Same |
| 2016-17 | 9 | 2/2/2020 | 0 | Same |
| 2017-18 | 9 | 1/19/2020 | -2 | Earlier |
| 2018-19 | 9 | 2/2/2020 | 0 | Same |

Leave-one-out detection date stability analysis, Prophet flat growth model, FluSurv-NET, United States. Each row reflects a model refit with one pre-pandemic season excluded from training (n = 9 seasons per run). First detection date is defined as the first post-training week in which observed rates exceeded the 95% prediction interval upper bound. Gap in weeks is calculated relative to the full-model detection date (2020-02-02). Detection date was identical to the full model in 9 of 10 leave-one-out runs. Excluding 2017–2018, the most anomalous pre-pandemic season, advanced detection by 2 weeks, reflecting a marginally lower training baseline in that run. No leave-one-out specification delayed detection, confirming that the February 2020 finding is not attributable to any single training season.

**Table S4C**. Cross-method convergence of Prophet forecast gaps and Isolation Forest anomaly classifications by season, FluSurv-NET, United States, 2009–2025.

| Season | Phase | Prophet Mean Gap | Prophet Gap Direction | IF Rank | IF Anomaly Score | IF Classification | Method Agreement |
|---|---|---|---|---|---|---|---|
| 2009-10 | PRE | -0.588 | Negative | 8 | -0.516 | Normal | Yes — both methods normal |
| 2010-11 | PRE | -0.801 | Negative | 9 | -0.453 | Normal | Yes — both methods normal |
| 2011-12 | PRE | -1.24 | Negative | 7 | -0.529 | Normal | Yes — both methods normal |
| 2012-13 | PRE | -0.062 | Negative | 12 | -0.433 | Normal | Yes — both methods normal |
| 2013-14 | PRE | -0.357 | Negative | 14 | -0.421 | Normal | Yes — both methods normal |
| 2014-15 | PRE | 0.398 | Positive | 4 | -0.569 | Normal | Partial — methods diverge |
| 2015-16 | PRE | -0.375 | Negative | 15 | -0.401 | Normal | Yes — both methods normal |
| 2016-17 | PRE | 0.541 | Positive | 13 | -0.428 | Normal | Partial — methods diverge |
| 2017-18 | PRE | 1.896 | Positive | 1 | -0.623 | Anomaly | Yes — both methods flag |
| 2018-19 | PRE | 0.588 | Positive | 10 | -0.447 | Normal | Partial — methods diverge |
| 2019-20 | DISR | 0.666 | Positive | 11 | -0.444 | Normal | Partial — methods diverge |
| 2021-22 | DISR | 1.352 | Positive | 6 | -0.538 | Normal | Partial — methods diverge |
| 2022-23 | REC | 0.557 | Positive | 5 | -0.553 | Normal | Partial — methods diverge |
| 2023-24 | REC | 0.541 | Positive | 3 | -0.6 | Anomaly | Yes — both methods flag |
| 2024-25 | REC | 2.68 | Positive | 2 | -0.615 | Anomaly | Yes — both methods flag |

Cross-method convergence: Prophet forecast gap direction and Isolation Forest classification by season, FluSurv-NET, United States, 2009–2025. Prophet gap direction indicates whether mean observed rates exceeded (Positive) or fell below (Negative) the flat growth model projection. Isolation Forest classification is from the primary model (contamination = 0.10). Method agreement is defined as concordance between a positive Prophet gap and an Isolation Forest anomaly classification, or between a negative gap and a normal classification. Partial divergence reflects the complementary scope of the two methods: Prophet measures deviation from a mean rate baseline while Isolation Forest detects multivariate extremeness across six seasonal features simultaneously. All three robust anomalies (2017–2018, 2023–2024, 2024–2025) show full agreement across both methods. The 2020–21 season was excluded from Isolation Forest scoring due to CDC structural data suppression.
